# Local genetic correlations exist among neurodegenerative and neuropsychiatric diseases

**DOI:** 10.1101/2022.05.30.22275781

**Authors:** Regina H. Reynolds, Aaron Z. Wagen, Frida Lona-Durazo, Sonja W. Scholz, Maryam Shoai, John Hardy, Sarah A. Gagliano Taliun, Mina Ryten

**Affiliations:** Genetics and Genomic Medicine, Great Ormond Street Institute of Child Health, University College London, London, UK; Aligning Science Across Parkinson’s (ASAP) Collaborative Research Network, Chevy Chase, MD 20815, United States; Department of Clinical and Movement Neurosciences, Queen Square Institute of Neurology, London, UK; Neurodegeneration Biology Laboratory, The Francis Crick Institute, London, UK; Montréal Heart Institute, Montréal, Québec, Canada; Neurodegenerative Diseases Research Unit, National Institute of Neurological Disorders and Stroke, Bethesda, MD, USA; Department of Neurology, Johns Hopkins University Medical Center, Baltimore, MD, USA; Department of Neurodegenerative Diseases, Queen Square Institute of Neurology, University College London, London, UK; Department of Medicine & Department of Neurosciences, Université de Montréal, Montréal, Québec, Canada; UK Dementia Research Institute, University College London, London, UK; NIHR Great Ormond Street Hospital Biomedical Research Centre, University College London, London, UK

## Abstract

Genetic correlation (*r*_*g*_) between traits can offer valuable insight into underlying shared biological mechanisms. Neurodegenerative diseases overlap neuropathologically and often manifest comorbid neuropsychiatric symptoms. However, global *r*_*g*_ analyses show minimal *r*_*g*_ among neurodegenerative and neuropsychiatric diseases. Importantly, local *r*_*g*_s can exist in the absence of global relationships. To investigate this possibility, we applied LAVA, a tool for local *r*_*g*_ analysis, to genome-wide association studies of 3 neurodegenerative diseases (Alzheimer’s disease, Lewy body dementia and Parkinson’s disease) and 3 neuropsychiatric disorders (bipolar disorder, major depressive disorder and schizophrenia). We identified several local *r*_*g*_s missed in global analyses, including between (i) all 3 neurodegenerative diseases and schizophrenia and (ii) Alzheimer’s and Parkinson’s disease. For those local *r*_*g*_s identified in genomic regions containing disease-implicated genes, such as *SNCA, CLU* and *APOE*, incorporation of expression quantitative trait loci identified genes that may drive genetic overlaps between diseases. Collectively, we demonstrate that complex genetic relationships exist among neurodegenerative and neuropsychiatric diseases, highlighting putative pleiotropic genomic regions and genes. These findings imply sharing of pathogenic processes and the potential existence of common therapeutic targets.

## Introduction

Neurodegenerative diseases are a group of syndromically-defined disorders that are characterised by the progressive loss of the structure and function of the central nervous system. They are typically grouped by their predominant neuropathological protein deposit (e.g. synucleinopathies, like Parkinson’s disease and Lewy body dementia, by α-synuclein deposition and Alzheimer’s disease by deposition of amyloid), but more often than not, they present with co-pathologies, suggesting that they might share common pathogenic pathways^1,2^. This notion is supported by genome-wide association studies (GWASs), which have (i) identified shared risk loci across neurodegenerative diseases, such as *APOE* and *BIN1* in Alzheimer’s disease (AD) and Lewy body dementia (LBD), or GBA, *SNCA*, TMEM175 in Parkinson’s disease (PD) and LBD and (ii) demonstrated that genetic risk scores derived from one neurodegenerative disease can predict risk of another, as with AD and PD scores predicting risk of LBD^3–5^. The importance of identifying common pathogenic processes cannot be overstated, given the implications for our mechanistic understanding of these diseases as well as identification of common therapeutic targets benefitting a wider range of patients.

From a clinical perspective, neurodegenerative diseases are often also defined in terms of their predominant symptom (e.g. AD by memory impairment or PD by parkinsonism), but in reality, present as highly heterogenous diseases, with symptoms spanning multiple domains including neuropsychiatric symptoms^6,7^. Indeed, a higher prevalence of depression has been observed in individuals with dementia compared to those without dementia^8^. Furthermore, depression and anxiety are more common in individuals with PD compared to the general population, with clinically significant symptoms in 30-35% of patients^9,10^. A similar (albeit reversed) phenomenon has been observed in some neuropsychiatric disorders, with a higher risk of dementia diagnoses observed in individuals with schizophrenia (SCZ) versus individuals without a history of serious mental illness^11,12^ and a higher risk of PD in individuals diagnosed with depressive disorder in mid or late life^10,13,14^. Together, these observations suggest the possibility of intersecting pathways between neurodegenerative and neuropsychiatric diseases.

Given these clinical and neuropathological overlaps, genetic overlaps would also be expected. However, a study of global genetic correlation between neurological phenotypes demonstrated limited overlap between individual neurodegenerative diseases as well as between neurodegenerative diseases and neuropsychiatric disorders^15,16^. Genetic correlation (*r*_*g*_) is a frequently used measure of genetic overlap, which is traditionally studied on a genome-wide scale, and thus, represents an average of the shared genetic effects across all causal loci in the genome^17^. This global approach may not capture shared genetic effects that are confined to particular regions of the genome (i.e. local *r*_*g*_s) or local *r*_*g*_s that have opposing directions across the genome^15,17^. Indeed, local *r*_*g*_s have been observed between neuropsychiatric traits^18^ and between AD and PD (specifically in the *HLA*^19^ and *MAPT* loci^20^). In addition, previous work using approaches that can detect polygenic overlap (including overlaps where there are mixed patterns of allelic effect directions) have demonstrated a global polygenic overlap between neurodegenerative and neuropsychiatric diseases, such as AD and BIP^21^, AD and MDD^22,23^, and PD and SCZ^24^. Collectively, these studies indicate that local genetic overlaps likely exist between neurodegenerative and neuropsychiatric diseases.

Here, we assess local *r*_*g*_ between 3 neurogenerative diseases (AD, LBD and PD) and 3 neuropsychiatric disorders (bipolar disorder, BIP; major depressive disorder, MDD; and SCZ). All 6 disease traits represent globally prevalent diseases^25^, have reasonably large GWAS cohorts ^3,5,26–30^, and importantly, have demonstrated evidence of a potential genetic overlap (but have not, to our knowledge, been systematically assessed all together for local *r*_*g*_). To estimate local *r*_*g*_ from GWAS summary statistics, we used the recently developed tool <u>l</u>ocal <u>a</u>nalysis of [co]<u>v</u>ariant <u>a</u>nnotation (LAVA)^31^. Unlike existing tools, such as rho-HESS^32^ and SUPERGNOVA^33^, which only permit testing of local *r*_*g*_s between two traits, LAVA is additionally able to model local genetic relations using more than two traits simultaneously, thus permitting exploration of local conditional genetic relations between multiple traits (a particularly useful feature in the context of neurodegenerative diseases like LBD, which has been hypothesised to lie on a disease continuum between AD and PD^5,34^). In addition, we use data from blood- and brain-derived gene expression traits, in the form of expression quantitative loci (eQTLs), to facilitate functional interpretation of local *r*_*g*_s between disease traits.

## Results

### Local analyses reveal genetic correlations among neurodegenerative and neuropsychiatric diseases

We applied LAVA to 3 neurodegenerative diseases (AD, LBD and PD) and 3 neuropsychiatric disorders (BIP, MDD and SCZ) (**Table 1**), all of which represent globally prevalent diseases^25^. Among neurodegenerative diseases, AD and PD are the most common, with a global prevalence of 8.98% and 1.12% in individuals > 70 years of age^6,25,35^ and consequently, have large GWAS cohorts (AD, N cases = 71,880; PD, N cases = 33,674)^3,26^. LBD is the second most common dementia subtype after AD, affecting between 4.2-30% of dementia patients^36^. As such, the LBD GWAS cohort is small (N cases = 2,591), but unlike AD and PD neurodegenerative GWASs, 69% of the cohort is pathologically defined^5^. Among neuropsychiatric disorders, MDD is the second most prevalent, with an estimated 185 million people affected globally (equivalent to 2.49% of the general population), while BIP and SCZ have a prevalence of 0.53% and 0.32%, respectively^25^. Accordingly, all 3 disorders have large, well-powered GWASs (BIP, N cases = 41,917; MDD, N cases = 170,756; SCZ, N cases = 40,675)^28–30^.

**Table 1.** Overview of traits included in this study. Global observed-scale SNP heritability (h^2^) for each trait computed using LDSC^86^ assuming a continuous liability (which may differ from liability scale h^2^ estimates). The h^2^ Z-score was calculated by dividing the global h^2^ estimate by its standard error (SE).

| Trait type | Trait | Abbreviation | N | N cases | N controls | Global $h^2$<br>(SE) | $h^2$ Z-score | Original study |
| --- | --- | --- | --- | --- | --- | --- | --- | --- |
| Disease | Alzheimer's disease<br>Clinically diagnosed + UK Biobank proxy cases and controls | AD | 455,258 | 71,880<br>(46,613 proxy) | 383,378<br>(318,246 proxy) | 1.5% (0.2) | 7.14 | Jansen *et al.*, 2019<sup>26</sup> |
| Disease | Alzheimer's disease<br>Clinically diagnosed | AD (no proxy) | 63,926 | 21,982 | 41,944 | 7.1% (1.1) | 6.25 | Kunkle *et al.*, 2019<sup>27</sup> |
| Disease | Bipolar disorder | BIP | 413,466 | 41,917 | 371,549 | 7.1% (0.3) | 26.2 | Mullins *et al.*, 2021<sup>28</sup> |
| Disease | Lewy body dementia<br>Autopsy-confirmed + clinically diagnosed | LBD | 6,618 | 2,591<br>(1,789 autopsy-confirmed) | 4,027 | 17.1% (7.6) | 2.27 | Chia *et al.*, 2021<sup>5</sup> |
| Disease | Major depressive disorder excluding 23andMe<br>Clinically diagnosed (33 PGC cohorts) + UK Biobank broad depression phenotype | MDD | 500,199 | 170,756 | 329,443 | 6% (0.2) | 26 | Howard *et al.*, 2019<sup>29</sup> |
| Disease | Parkinson's disease excluding 23andMe<br>Clinically diagnosed + UK Biobank proxy cases and controls | PD | 482,730 | 33,674<br>(18,618 proxy) | 449,056<br>(436,419 proxy) | 1.9% (0.2) | 10.9 | Nalls *et al.*, 2019<sup>3</sup> |
| Disease | Parkinson's disease excluding 23andMe<br>Clinically diagnosed | PD (no proxy) | 27,693 | 15,056 | 12,637 | 30.6% (2.8) | 11.1 | Nalls *et al.*, 2019<sup>3</sup> |
| Disease | Schizophrenia | SCZ | 105,318 | 40,675 | 64,643 | 41% (1.4) | 29.7 | Pardiñas *et al.*, 2018<sup>30</sup> |
| Gene expression | eQTLGen<br>Blood-derived eQTLs | eQTLGEN | 31,684 | - | - | - |  | Vosã *et al.*, 2021<sup>43</sup> |
| Gene expression | PsychENCODE<br>Brain-derived eQTLs | PSYCHENCODE | 1,387 | - | - | - |  | Wang *et al.*, 2018<sup>44</sup> |

We tested pairwise local genetic correlations (*r*_*g*_s) across a targeted subset of 300 local autosomal genomic regions that contain genome-wide significant GWAS loci from at least one trait (**Supplementary Figure 1, Supplementary Table 1**). These genomic regions, henceforth referred to as linkage disequilibrium (LD) blocks, were filtered from the original 2,495 LD blocks generated by Werme *et al*.^31^ using a genome-wide partitioning algorithm that aims to reduce LD between LD blocks.

First, we performed a univariate test for every disease trait at each of the 300 LD blocks to ensure sufficient local genetic signal was present to proceed with bivariate local *r*_*g*_ analyses. Pairs of traits exhibiting a univariate local genetic signal of p < 0.05/300 were then carried forward to bivariate tests, resulting in 1,603 bivariate tests across 275 distinct LD blocks. Using a Bonferroni-corrected p-value threshold of p < 0.05/1,603, we detected 77 significant bivariate local *r*_*g*_s across 59 distinct LD blocks, with 25 local *r*_*g*_s between trait pairs where no significant global *r*_*g*_ was observed (**Figure 1a, Figure 1b, Supplementary Table 2, Supplementary Table 3**). These 25 correlations included: (i) local *r*_*g*_s between all 3 neurodegenerative diseases and SCZ; (ii) a local *r*_*g*_ between PD and BIP; and (ii) 20 local *r*_*g*_s between AD and PD.For 30 of the 77 local *r*_*g*_s, the genetic signal of both disease traits may overlap entirely, suggested by the upper limit of the 95% confidence interval (CI) for explained variance (i.e. *r*^2^, the proportion of variance in genetic signal of one disease trait in a pair explained by the other) including 1. Notably, the trait pairs where the upper limit of the 95% CI did not include 1 all involved at least one neurodegenerative disease, with the one exception being a local *r*_*g*_ between PD and SCZ, suggesting that the genetic overlap between neurodegenerative diseases is smaller than between neuropsychiatric disorders in the tested LD blocks (**Figure 1c**).

**Figure 1.**
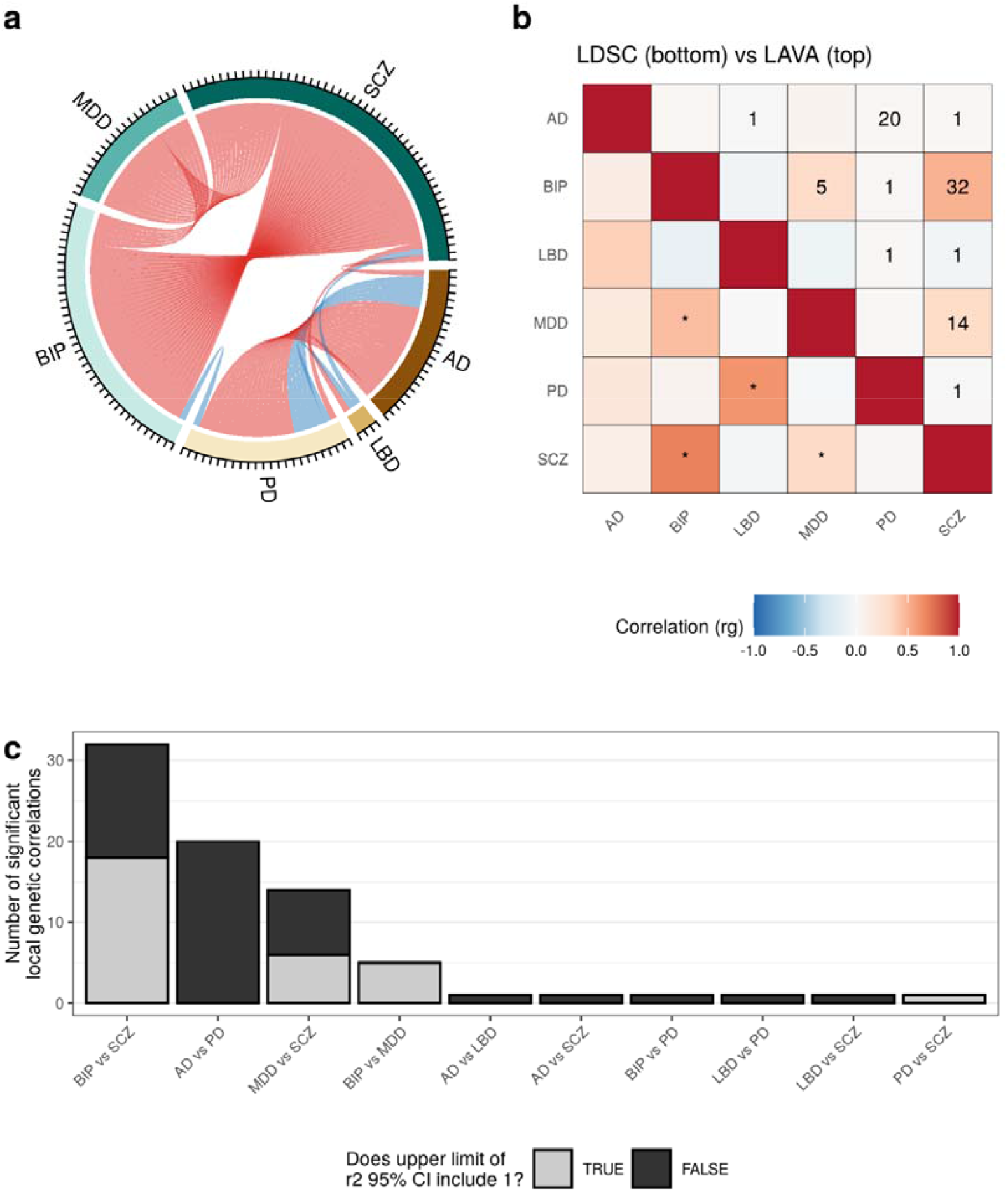
Overview of local and global genetic correlations between neurodegenerative diseases and neuropsychiatric disorders. **(a)** Chord diagram showing the number of significant bivariate local s (p < 0.05/1603) between each of the disease traits across all LD blocks. Positive and negative correlations are coloured red and blue, respectively. **(b)** Comparison between the global s estimated by LDSC (bottom) and the mean local from LAVA (top) across all tested LD blocks. Significant global s (p < 0.05/15) are indicated with *. The number of significant local s is indicated by a number in each tile. **(c)** Bar plot showing the number of significant local s between disease trait pairs. The fill of the bars indicates the number of significant LD blocks for which the upper limit of the 95% confidence interval (CI) included 1.

We intersected local *r*_*g*_s with previously reported local genetic associations, which were derived using tools for local *r*_*g*_ estimation (rho-HESS^19^ and LAVA^18^) and estimation of global polygenic overlap (the conditional/conjunctional FDR approach^23,24,37^). We found no overlap between local *r*_*g*_s from our study and local genetic associations reported using rho-HESS (which included an association between AD and PD in the *HLA* locus) or the conditional/conjunctional FDR approach (**Supplementary Note, Supplementary Table 4**). We did, however, demonstrate an overlap between local *r*_*g*_s from our study and local *r*_*g*_s reported by Gerring *et al*., which also used LAVA, albeit to study 10 psychiatric disorders and 10 substance abuse phenotypes. Between the two studies, bivariate tests for the same trait pairs were performed in 6 LD blocks. We were able to replicate 5 of the 7 overlapping local *r*_*g*_s (BIP and SCZ in LD block 457; SCZ and BIP or MDD in LD block 951; MDD and SCZ in LD block 952; and BIP and SCZ in LD block 2483; **Supplementary Note; Supplementary Figure 2; Supplementary Table 4**).

### Local analyses associate disease-implicated genomic regions with previously unrelated traits

Across the 77 local *r*_*g*_s, 22 involved trait pairs where both traits had genome-wide significant single nucleotide polymorphisms (SNPs) overlapping the LD block tested, 35 involved trait pairs where one trait in the pair had genome-wide significant SNPs overlapping the LD block tested and 20 involved trait pairs where neither trait had genome-wide significant SNPs overlapping the LD block tested (**Figure 2a**). Thus, despite the targeted nature of our approach (which biased analyses towards LD blocks that contain genome-wide significant GWAS SNPs), 71% of the detected local *r*_*g*_s linked genomic regions implicated by one of the six disease traits with seemingly unrelated disease traits. For example, LD block 1719 (chr11:112,755,447-113,889,019) and 2281 (chr18:52,512,524-53,762,996) both contained genome-wide significant GWAS SNPs from MDD and SCZ, an overlap which was mirrored by a significant local *r*_*g*_ between MDD and SCZ (**Figure 2b**). In addition, both LD blocks implicated disease traits that did not have overlapping genome-wide significant GWAS SNPs in the region, indicating novel disease trait associations. These included (i) LBD in LD block 1719 (chr11:112,755,447-113,889,019), which negatively correlated with SCZ (ρ = -0.65, p = 4.72 × 10^−6^) and (ii) AD and PD, which were positively correlated in LD block 2281 (chr18:52,512,524-53,762,996, ρ = 0.41, p = 1.24 × 10^−8^). Notably, both LD blocks contain genes of interest to traits implicated by local *r*_*g*_ analyses, including *DRD2* in LD block 1719 (encodes dopamine receptor D2, a target of drugs used in both PD^7^ and SCZ treatment^38^) and RAB27B in LD block 2281 (encodes Rab27b, a Rab GTPase recently implicated in α-synuclein clearance^39^).

**Figure 2.**
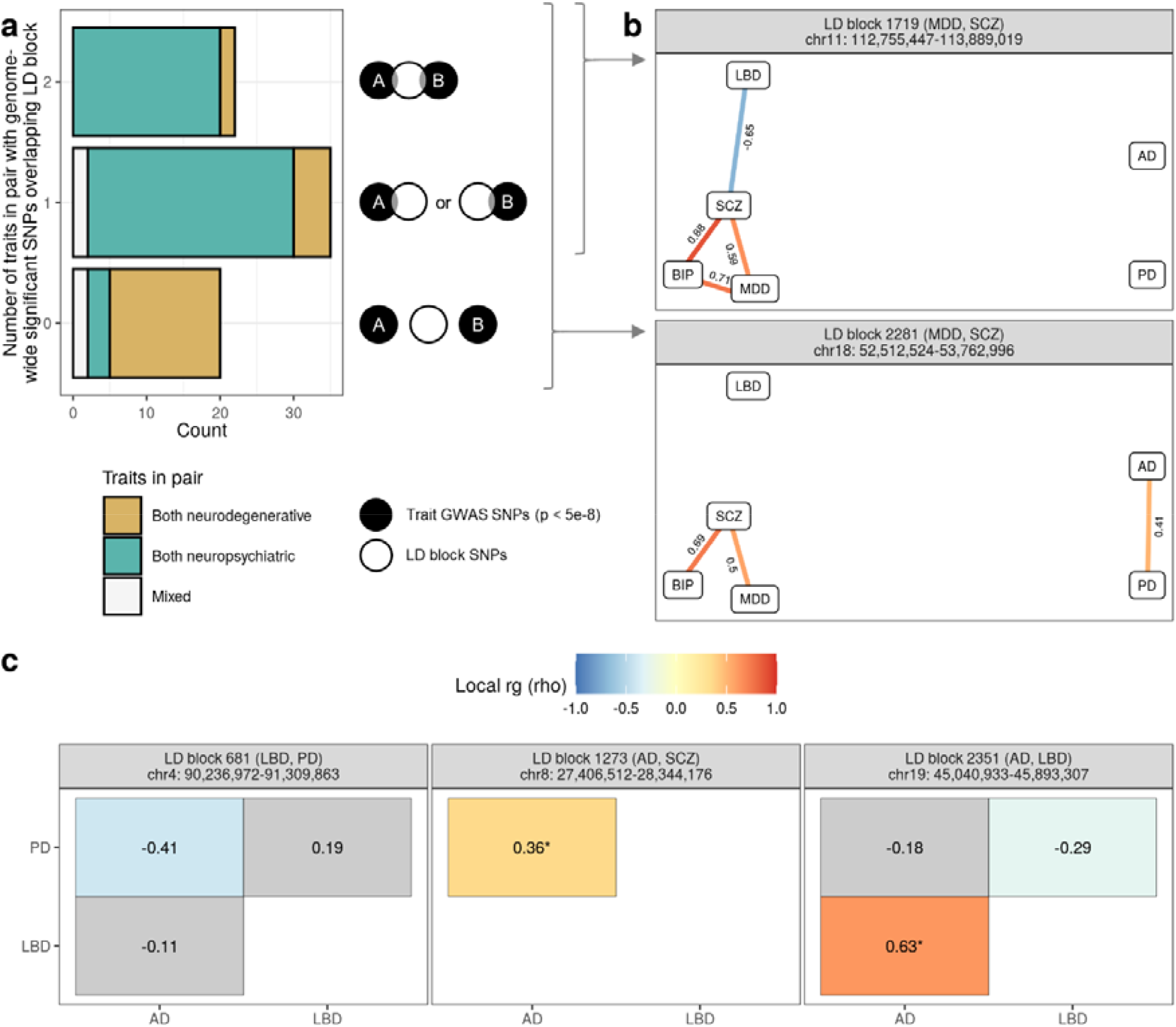
Local analyses associate disease-implicated genomic regions with previously unrelated traits. **(a)** Bar plot (left) showing the number of traits within trait pairs demonstrating significant local s that had genome-wide significant SNPs overlapping the tested LD block (as illustrated by the schematic on the right**). (b)** Two LD blocks illustrating the situations depicted in **(a)**. Edge diagrams for each LD block show the standardised coefficient for (rho, ρ) for each significant bivariate local. Significant negative and positive s are indicated by blue and red colour, respectively. **(c**) Heatmaps show the rho for each bivariate local within the LD block. Asterisks (*) indicate s that were replicated when using AD and PD GWASs that excluded UK Biobank by-proxy cases. Significant negative and positive s are indicated by blue and red fill, respectively. Non-significant s have a grey fill. In both **(b)** and **(c)** panels are labelled by the LD block identifier, the traits with genome-wide significant SNPs overlapping the LD block (indicated in the brackets) and the genomic coordinates of the LD block (in the format chromosome:start-end, GRCh37).

Local *r*_*g*_ analyses also highlighted relationships between neurodegenerative traits in regions containing well-known, disease-implicated genes, such as: (i) *SNCA* (implicated in monogenic and sporadic forms of PD^3,5^) in LD block 681 (chr4:90,236,972-91,309,863), where a negative local *r*_*g*_ was observed between AD and PD (ρ = -0.41, p = 6.51 × 10^−13^); (ii) *CLU* (associated with sporadic AD^26,40^) in LD block 1273 (chr8:27,406,512-28,344,176), where a positive local *r*_*g*_ was observed between AD and PD (ρ = 0.36, p = 8.76 × 10^−12^); and finally, (iii) *APOE* (E4 alleles associated with increased AD risk^41^) in LD block 2351 (chr19:45,040,933-45,893,307), where *r*_*g*_s were observed between LBD and both AD and PD (LBD-AD: ρ = 0.59, p = 1.24 × 10^−139^; LBD-PD: ρ = -0.29, p = 2.75 × 10^−7^) (**Figure 2c**). We also noted a positive correlation between AD and PD in LD block 2128 (chr16:29,043,178-31,384,210), which contains the AD-associated *KAT8* locus^26^ and the PD-associated *SETD1A* locus^3^ (of note, rare loss-of-function variants in *SETD1A* are associated with schizophrenia^42^).

### Sensitivity analysis indicates that by-proxy cases do not drive spurious local correlations among neurodegenerative diseases

Given concerns that UK Biobank (UKBB) by-proxy cases could potentially be mislabelled (i.e. parents of by-proxy case suffered from another type of dementia) and lead to spurious *r*_*g*_s between neurodegenerative traits, we performed sensitivity analyses using GWASs for AD and PD that excluded UKBB by-proxy cases^27^. Of the 21 LD blocks where significant local *r*_*g*_s were observed between the 3 neurodegenerative traits using AD and PD GWASs with by-proxy cases, only LD block 1273 (chr8:27,406,512-28,344,176) and LD block 2351 (chr19:45,040,933-45,893,307) had sufficient local genetic signal for both AD and PD without by-proxy cases. This likely reflects the decrease in cohort numbers when UKBB by-proxy cases are excluded from AD and PD GWASs (**Table 1**). We were able to replicate 2 of the 3 significant local *r*_*g*_s observed in LD block 1273 (chr8:27,406,512-28,344,176) and 2351 (chr19:45,040,933-45,893,307), including the positive *r*_*g*_ between AD and PD in LD block 1273 and the positive *r*_*g*_ between AD and LBD in LD block 2351 (**Supplementary Figure 3, Supplementary Table 5**). Further, while the local *r*_*g*_ between LBD and PD in LD block 2351 was non-significant when using the PD GWAS without by-proxy cases, the correlation was in the same direction in the complementary analysis using the PD GWAS with by-proxy cases (no by-proxy: ρ = - 0.201, ρ CI = -0.443 to 0.007, p = 0.061; by-proxy: ρ = -0.293, ρ CI = -0.405 to -0.184, p = 2.75 × 10^−7^) (**Supplementary Figure 3, Supplementary Table 5**).

### Local heritability of Lewy body dementia in an *APOE-*containing LD block is only partly explained by Alzheimer’s disease and Parkinson’s disease

Eleven LD blocks were associated with > 1 trait pair, of which 8 LD blocks had a trait in common across multiple trait pairs. In other words, the genetic component of one disease trait (the outcome trait) could be modelled using the genetic components of multiple predictor disease traits. This included 3 LD blocks (LD block 758, chr4:175,959,698-177,129,678; LD block 951, chr6:26,396,201-27,261,035; and LD block 952, chr6:27,261,036-28,666,364) where all 3 neuropsychiatric disorders were significantly correlated with one another, and thus could arguably be the outcome trait. In these situations, each neuropsychiatric disorder was separately modelled as the outcome trait, resulting in 3 independent models within each of these 3 LD blocks. The remaining 5 LD blocks only had 1 trait in common across correlated trait pairs, therefore only one model was constructed for each. A total of 14 multivariate models were run across all 8 LD blocks, of which 6 models were found to contain a predictor trait that significantly contributed to the local heritability of an outcome trait (**Figure 3a, Supplementary Table 6**).

**Figure 3.**
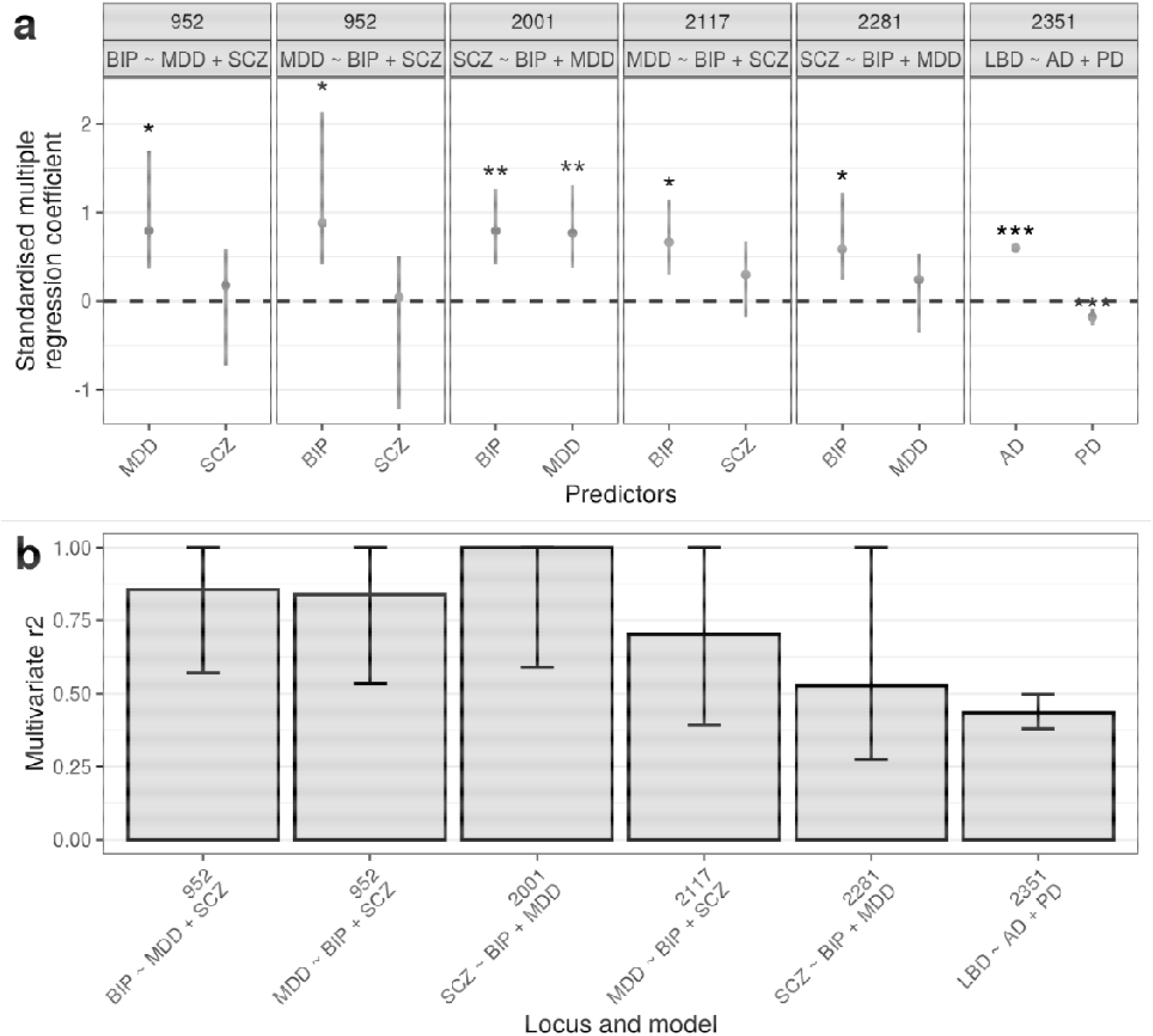
Multiple regression across LD blocks with multiple trait pair correlations. For both plots, only those multiple regression models with at least one significant predictor (p < 0.05) are shown. **(a)** Plots of standardised coefficients for each predictor in multiple regression models across each LD block, with whiskers spanning the 95% confidence interval for the coefficients. Panels are labelled by the LD block identifier and the regression model. **(b)** Multivariate for each LD block and model, where multivariate represents the proportion of variance in genetic signal for the outcome trait explained by all predictor traits simultaneously. Whiskers span the 95% confidence interval for the. ***, p < 0.001; **, p < 0.01; *, p < 0.05. Coordinates for LD blocks (in the format chromosome:start-end, GRCh37): 952, chr6:27,261,036-28,666,364; 2001, chr14:71,140,427-72,665,319; 2117, chr16:12,793,150-13,893,407; 2281, chr18:52,512,524-53,762,996; 2351, chr19:45,040,933-45,893,307.

We noted that all models with a neuropsychiatric outcome trait and a significant neuropsychiatric predictor trait had a high multivariate *r*^2^ (range: 0.53-1), with upper confidence intervals including 1 (**Figure 3b**), suggesting that the genetic signal of the neuropsychiatric outcome trait could be entirely explained by its predictor traits in these LD blocks. In contrast, in the *APOE*-containing LD block 2351 (chr19:45,040,933-45,893,307), which was modelled with LBD as the outcome and AD and PD as predictors, the multivariate *r*^2^ was 0.43 (95% CI: 0.38 to 0.5), a result that held using GWASs for AD and PD that excluded by-proxy cases (*r*^2^ = 0.49, 95% CI: 0.44 to 0.57; **Supplementary Figure 3**). Thus, while AD and PD jointly explained approximately 40% of the local heritability of LBD, a proportion of the local heritability for LBD was independent of AD and PD.

### Incorporation of gene expression traits to facilitate functional interpretation of disease trait correlations

To dissect whether regulation of gene expression might underlie local *r*_*g*_s between disease traits, we performed local *r*_*g*_ analyses using expression quantitative trait loci (eQTLs) from eQTLGen^43^ and PsychENCODE^44^, which represent large human blood and brain expression datasets, respectively (**Table 1**). We used LAVA to study relationships between gene expression and disease traits on account of its ability to model the uncertainty in eQTL effect estimates (unlike the commonly used TWAS framework, which has been shown to have an increased type 1 error rate^45^, as a result of it not accounting for the uncertainty in the estimated genetic component of gene expression). In addition, where three-way relationships were observed between 2 disease traits and an eQTL, we computed partial correlations to determine whether correlations between disease traits could be explained by the eQTL.

We restricted analyses to the 5 LD blocks highlighted in **Figure 2** (LD block 681, chr4:90,236,972-91,309,863; LD block 1273, chr8:27,406,512-28,344,176; LD block 1719, chr11:112,755,447-113,889,019; LD block 2281, chr18:52,512,524-53,762,996; LD block 2351, chr19:45,040,933-45,893,307), which contained genes of interest to at least one of the disease traits implicated by local *r*_*g*_ analyses. From these LD blocks of interest, we defined genic regions (gene start and end coordinates ± 100 kb) for all overlapping protein-coding, antisense or lincRNA genes (*n* = 92).

We detected a total of 135 significant bivariate local *r*_*g*_s across 47 distinct genic regions (FDR < 0.05), with 43 local *r*_*g*_s across 27 distinct genic regions between trait pairs involving a disease trait and a gene expression trait (**Supplementary Figure 4, Supplementary Table 7**). We noted that the explained variance (*r*^2^) between trait pairs involving a disease trait and a gene expression trait tended to be lower than between trait pairs involving two disease traits **(Supplementary Figure 5**), an observation that aligns with a recent study that found only 11% of trait heritability to be mediated by bulk-tissue gene expression^46^.

With the exception of the *SNCA*-containing LD block 681 (chr4:90,236,972-91,309,863), where eQTLs for only 1 out of 5 genes tested in the block were correlated with a disease trait (negative *r*_*g*_ between blood-derived *SNCA* eQTLs and PD), the expression of multiple genes was associated with disease traits across the remaining LD blocks (**Figure 4a**). In addition, the expression of several genes was associated with more than one disease trait (**Figure 4b**). For example, blood- and brain-derived *ANKK1* eQTLs (*DRD2*-containing LD block 1719, chr11:112,755,447-113,889,019) were negatively correlated with both MDD and SCZ, which themselves were positively correlated (**Figure 4c**). A SNP residing in the coding region of *ANKK1* (rs1800497, commonly known as TaqIA SNP) has been previously associated with alcoholism, schizophrenia and eating disorders, although it is unclear whether this SNP exerts its effect via *DRD2* or *ANKK1*^47^. As *DRD2* is not expressed in blood, and brain-derived *DRD2* eQTLs did not pass the univariate test for sufficient local genetic signal, we were unable to test for local *r*_*g*_s between *DRD2* eQTLs and any neuropsychiatric disorder. The data available would therefore suggest that the shared risk of MDD and SCZ in the overlapping *ANKK1* and *DRD2* genic regions may be partly driven by *ANKK1* gene expression. Indeed, the local *r*_*g*_ between MDD and SCZ was weakened when conditioned on *ANKK1* eQTLs (eQTLGen: MDD∼SCZ, *r*_*g*_ = 0.72, p = 0.000132; MDD∼SCZ|*ANKK1, r*_*g*_ = 0.60, p = 0.0203; PsychENCODE: MDD∼SCZ, *r*_*g*_ = 0.67, p = 0.000271; MDD∼SCZ|*ANKK1, r*_*g*_ = 0.61, p = 0.00441; **Supplementary Table 9**).

**Figure 4.**
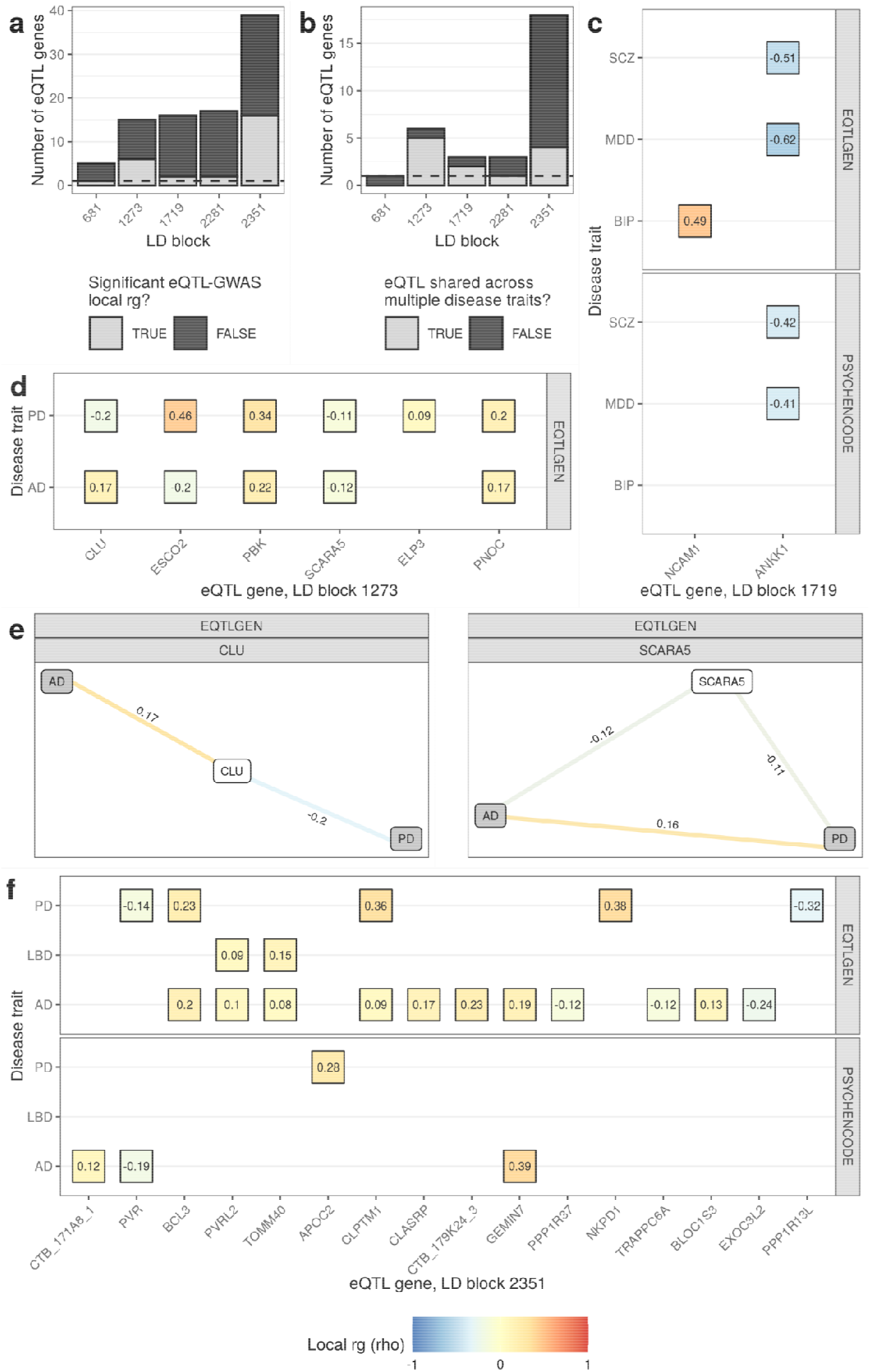
Incorporation of gene expression traits to facilitate functional interpretation of disease trait correlations. **(a)** Bar plot of the number of eQTL genes (as defined by their genic regions) tested in each LD block. The fill of the bars indicates whether eQTL genes were significantly correlated with at least one disease trait. **(b)** Bar plot of the number of eQTL genes that were significantly correlated with at least one disease trait. The fill of the bars indicates whether eQTL genes in local *r*_*g*_s were correlated with one or more disease traits. **(c, d, f)** Heatmaps of the standardised coefficient for *r*_*g*_ (rho) for each significant gene expression-disease trait correlation (FDR < 0.05) within LD block (c) 1719, **(d)** 1273 and **(f)** 2351. Genes are ordered left to right on the x-axis by the genomic coordinate of their gene start. Panels are labelled by the eQTL dataset from which eQTL genes were derived (either PsychENCODE’s analysis of adult brain tissue from 1387 individuals or the eQTLGen meta-analysis of 31,684 blood samples from 37 cohorts**). (e)** Edge diagrams for representative genic regions show the rho for each significant bivariate local *r*_*g*_ (FDR < 0.05). GWAS and eQTL nodes are indicated by grey and white fill, respectively. Panels are labelled by the gene tested and the eQTL dataset from which eQTL genes were derived. In panels c-f significant negative and positive *r*_*g*_s are indicated by blue and red colour, respectively. Coordinates for LD blocks (in the format chromosome:start-end, GRCh37): 681, chr4:90,236,972-91,309,863; 1273, chr8:27,406,512-28,344,176; 1719, chr11:112,755,447-113,889,019; 2281, chr18:52,512,524-53,762,996; 2351, chr19:45,040,933-45,893,307.

A high degree of eQTL sharing across disease traits was observed in the *CLU*-containing LD block 1273 (chr8:27,406,512-28,344,176), with blood-derived eQTLs from 5 out of the 6 genes implicated in local *r*_*g*_s found to correlate with both AD and PD (**Figure 4b,d**). This included situations where eQTL-disease trait correlations had (i) the same direction of effect across both disease traits (as observed with *PBK, PNOC* and *SCARA5*) or (ii) opposing directions of effect across both disease traits (as observed with *CLU* and *ESCO2*) (**Figure 4d**). Notably, while a significant positive local *r*_*g*_ was observed between AD and PD in the *PBK* and *SCARA5* genic regions (reflecting the positive local *r*_*g*_ observed between AD and PD across the entire LD block), no local *r*_*g*_ was observed between AD and PD in the *CLU* genic region, suggesting that the shared risk of AD and PD in LD block 1273 may be driven by the expression of genes other than the AD-associated *CLU* (**Figure 4e**). As a ferritin receptor involved in ferritin internalisation, *SCARA5* could plausibly drive shared AD and PD risk, given that cellular iron overload and iron-induced oxidative stress have been implicated in several neurodegenerative diseases such as AD and PD^48,49^. However, conditioning the local *r*_*g*_ between AD and PD on *SCARA5* eQTLs had little effect on the strength and significance of the correlation (AD∼PD, *r*_*g*_ = 0.16, p = 0.000135; AD∼PD |*SCARA5, r*_*g*_ = 0.15, p = 0.000487**; Supplementary Table 9**), suggesting that the relationship of *SCARA5* eQTLs with AD and PD operates independently across each disease. In contrast, the local *r*_*g*_ between AD and PD was weakened and no longer significant after conditioning on *PBK* eQTLs (AD∼PD, *r*_*g*_ = 0.14, p = 0.0187; AD∼PD |*PBK, r*_*g*_ = 0.07, p = 0.259; **Supplementary Table 9**), indicating the *PBK* eQTLs may partly explain the local *r*_*g*_ between AD and PD. *PBK* encodes a serine-threonine kinase involved in regulation of cellular proliferation and cell-cycle progression^50^, which has been shown to be overexpressed in proliferative cells, including neural precursors cells in the subventricular zone of the adult brain^50,51^.

Compared to LD block 1273, the degree of eQTL sharing across disease traits was lower in the *APOE*-containing LD block 2351 (chr19:45,040,933-45,893,307), with eQTLs from 4 out of 16 genes implicated in local *r*_*g*_s found to correlate with AD and one of PD or LBD (**Figure 4b, f**). Shared eQTL genes were only observed in blood and included *BCL3, CLPTM1, PVRL2* and *TOMM40*, with expression of *BCL3* and *CLPTM1* positively correlating with AD and PD and expression of *PVRL2* and *TOMM40* positively correlating with AD and LBD. As the exception, *PVR* eQTLs were negatively associated with both AD and PD albeit in different tissues: AD in brain and PD in blood. Expression of the remaining 11 genes was exclusively associated with either AD (n = 8) or PD (n = 3). No significant local *r*_*g*_ was observed between *APOE* eQTLs and AD (FDR < 0.05), although a nominal positive *r*_*g*_ was observed in blood (ρ = 0.178, ρ CI = 0.007 to 0.352, p = 0.039; **Supplementary Figure 4e, Supplementary Table 7**). Overall, these results indicate that risk of neurodegenerative diseases (in particular, AD) is associated with expression of multiple genes in the *APOE*-containing LD block.

Further, they add to a growing body of evidence suggesting that in parallel with the well-studied *APOE-ε4* risk allele, there are additional *APOE*-independent risk factors in the region (such as *BCL3*^52^ and *PVRL2*^53^) that may contribute to AD risk.

For a complete overview of all genic regions tested across the 5 LD blocks of interest, see **Supplementary Figure 4 and Supplementary Table 7**.

## Discussion

Despite clinical and neuropathological overlaps between neurodegenerative diseases, global analyses of genetic correlation (*r*_*g*_) show minimal *r*_*g*_ among neurodegenerative diseases or across neurodegenerative and neuropsychiatric diseases. However, local *r*_*g*_s can deviate from the genome-wide average estimated by global analyses and may even exist in the absence of a genome-wide *r*_*g*_, thus motivating the use of tools to model local genetic relations.

Here, we applied LAVA to 3 neurodegenerative diseases and 3 neuropsychiatric disorders to determine whether local *r*_*g*_s exist in a subset of 300 LD blocks that contain genome-wide significant GWAS loci from at least one of six investigated disease traits. We identified 77 significant bivariate local *r*_*g*_s across 59 distinct LD blocks, with 25 local *r*_*g*_s between trait pairs where no significant global *r*_*g*_ was observed, including between (i) all 3 neurodegenerative diseases and SCZ and (ii) AD and PD. Local *r*_*g*_s highlighted expected associations (e.g. AD and LBD in the *APOE*-containing LD block 2351^5^, chr19:45,040,933-45,893,307) and putative new associations (e.g. AD and PD in the *CLU*-containing LD block 1273, chr8:27,406,512-28,344,176) in genomic regions containing well-known, disease-implicated genes. Likewise, incorporation of eQTLs confirmed known relationships between diseases and genes, such as the association of AD with *CLU* expression^40^ and PD with *SNCA* expression in blood^54^, and revealed putative new disease-gene relationships. Together, these results indicate that more complex aetiological relationships exist between neurodegenerative and neuropsychiatric diseases than those revealed by global *r*_*g*_s. Further, they highlight potential gene expression intermediaries that may account for local *r*_*g*_s between disease traits.

These findings have important implications for our understanding of neurodegenerative diseases and the extent to which they overlap. An overlap between the synucleinopathies and AD is often acknowledged in the context of LBD, which has been hypothesised to lie on a disease continuum between AD and PD^5,34^. In support of this continuum, LBD was found to associate with both AD and PD in the *APOE*-containing LD block 2351 (chr19:45,040,933-45,893,307). Multiple regression analyses confirmed that AD and PD were significant predictors of LBD heritability in LD block 2351. Importantly, when AD and PD were modelled together, they explained only ∼ 40% of the local heritability of LBD in LD block 2351, implying that LBD represents more than the union of AD and PD. Further, the associations of AD and PD with LBD had opposing regression coefficients, suggesting that the contribution of AD and PD to LBD in the *APOE* locus may not be synergistic. This mirrors the observation that genome-wide genetic risk scores of AD and PD do not interact in LBD,^5^ and may indicate that different biological pathways underlie the association between LBD and AD/PD. Indeed, only blood-derived *PVRL2* and *TOMM40* eQTLs were found to correlate with both AD and LBD, while no shared eQTL genes were detected between PD and LBD.

Less acknowledged is the genetic overlap between AD and PD, with no global *r*_*g*_ reported between the two diseases^16,55^ and no significant evidence for the presence of loci that increase the risk of both diseases^56^. As the exception, genetic overlaps have been reported between AD and PD in the *HLA*^19^ and *MAPT* loci^20^, hinting that pleiotropy may exist locally. In support of local pleiotropy, we observed 20 local *r*_*g*_s between AD and PD in genomic regions containing disease-implicated genes, such as *SNCA* (LD block 681, chr4:90,236,972-91,309,863) and *CLU* (LD block 1273, chr8:27,406,512-28,344,176). In the case of the *CLU*-containing LD block 1273 (chr8:27,406,512-28,344,176), incorporation of eQTLs demonstrated an association of AD and PD with the expression of 5 genes, although partial correlations suggested that only *PBK* expression could explain the correlation between AD and PD, with the remaining associations between eQTLs and AD or PD appearing to operate independently across diseases. In contrast, only blood-derived *SNCA* eQTLs were associated with PD in LD block 681 (chr4:90,236,972-91,309,863), suggesting that the association between AD and PD at the *SNCA* locus could be driven by tissue- or context-dependent gene expression or alternatively other molecular phenotypes.

A few studies have demonstrated genetic overlaps between neurodegenerative and neuropsychiatric diseases, such as AD and BIP^21^, AD and MDD^22,23^, and PD and SCZ^24^, while others have demonstrated no overlap^16,57^, with divergences in outcomes ascribed to differences in methodology and cohort^22^. Here, we observed a local *r*_*g*_ between BIP and PD, in addition to local *r*_*g*_s between schizophrenia and all 3 neurodegenerative diseases, which in the case of LBD was observed in an LD block containing the gene *DRD2* (LD block 1719, chr11:112,755,447-113,889,019). Notably, parkinsonism in dementia with Lewy bodies (DLB, one of the two LBDs), is often less responsive to dopaminergic treatments than in PD^58^. Furthermore, methylation of the *DRD2* promoter in leukocytes has been shown to differ between DLB and PD^59^, while D2 receptor density has been shown to be significantly reduced in the temporal cortex of DLB patients, but not AD^60^, suggesting that the *DRD2* locus may harbour markers that could distinguish between these neurodegenerative diseases. Our study adds to the body of evidence in favour of a shared genetic basis between neurodegenerative and neuropsychiatric diseases, although further work will be required to determine whether this genetic overlap underlies the clinical and epidemiological links observed between these two disease groups.

This study is not without its limitations, with several limitations related to the input data. These limitations include: (i) the variability in cohort size (sample size is a key determinant of the power to detect the association of a variant with a trait), which in the case of the smallest GWAS, LBD, may explain the limited number of local *r*_*g*_s observed involving this trait; (ii) the risk of misdiagnosis (particularly in GWASs that include broader definitions of a disorder, such as the MDD GWAS, which includes the UK Biobank broad definition of depression as well as clinically-derived phenotypes for MDD); and (iii) the lack of genetic diversity (i.e. all traits used were derived from individuals of European ancestry). Given that population-specific genetic risk factors exist, such as the lack of *MAPT* GWAS signal in the largest GWAS of Asian patients with PD^61^, and that transethnic global *r*_*g*_s between traits such as gene expression are significantly less than 1^62^, it is imperative that studies of local *r*_*g*_ are expanded to include diverse populations.

Among methodological limitations, both LDSC and LAVA only consider autosomal chromosomes, leaving out chromosome X, which is not only longer than chromosome 8-22, but also encodes 858 and 689 protein-coding and non-coding genes, respectively (Ensembl v106)^63^. Furthermore, as mentioned by the developers of LAVA^31^, local *r*_*g*_s could potentially be confounded by association signals from adjacent genomic regions, a limitation which is particularly pertinent in our analysis of gene expression traits where LD blocks were divided into smaller (often overlapping) genic regions. Additional fine-mapping (both computational and biological) could be helpful in narrowing down the set of potentially causal variants and consequently the genomic regions of interest^64^.

Importantly, as with any genetic correlation analysis, an observed *r*_*g*_ does not guarantee the presence of true pleiotropy. Spurious *r*_*g*_s can occur due to LD or misclassification^17^. Here, we attempted to address the potential misclassification of by-proxy cases via sensitivity analyses using GWASs for AD and PD that excluded UKBB by-proxy cases. We replicated 2 of the 3 significant local *r*_*g*_s observed in 2 LD blocks when using GWASs with by-proxy cases. However, we were unable to test for local *r*_*g*_s across the remaining 19 LD blocks due to insufficient univariate signal, which could reflect (i) a genuine contribution of by-proxy cases to trait *h*^2^ in the region or (ii) a lack of statistical power to detect a genetic signal. Given the substantial decrease in cohort numbers when UKBB by-proxy cases are excluded from AD and PD GWASs (**Table 1)**, a lack of statistical power seems the more likely explanation, warranting a revisit of this analysis as clinically-diagnosed and/or pathologically-defined cohorts increase in size.

Finally, even where observed *r*_*g*_s potentially represent true pleiotropy, LAVA cannot discriminate between vertical and horizontal pleiotropy (also known as mediated and biological pleiotropy, respectively^17,31^). Thus, while incorporation of gene expression can provide testable hypotheses regarding the underlying genes and biological pathways that drive relationships between neurodegenerative and neuropsychiatric diseases, experimental validation is required to establish the extent to which these genes represent genuine intermediary phenotypes.

In summary, our results have important implications for our understanding of the genetic architecture of neurodegenerative and neuropsychiatric diseases, including the demonstration of local pleiotropy particularly between neurodegenerative diseases. Not only do these findings suggest that neurodegenerative diseases may share common pathogenic processes, highlighting putative gene expression intermediaries which may underlie relationships between diseases, but they also infer the existence of common therapeutic targets across neurodegenerative diseases that could be leveraged for the benefit of broader patient groups.

## Materials and methods

### Trait pre-processing

Summary statistics from a total of 8 distinct traits were used, including 6 disease traits and 2 gene expression traits. Disease traits included 3 neurodegenerative diseases (Alzheimer’s disease, AD; Lewy body dementia, LBD; and Parkinson’s disease, PD) and 3 neuropsychiatric disorders (bipolar disorder, BIP; major depressive disorder, MDD; and schizophrenia, SCZ)^3,5,26–30^. Gene expression traits were used to facilitate functional interpretation of local genetic correlations (*r*_*g*_) between disease traits. Gene expression traits included expression quantitative trait loci (eQTLs) from eQTLGen^43^ and PsychENCODE^44^, which represent large human blood and brain expression datasets, respectively. All traits used were derived from individuals of European ancestry. Details of all summary statistics used can be found in **Table 1**.

Where necessary, SNP genomic coordinates were mapped to Reference SNP cluster IDs (rsIDs) using the SNPlocs.Hsapiens.dbSNP144.GRCh37 package^65^. In the case of the PD GWAS without UK Biobank (UKBB) data (summary statistics were kindly provided by the International Parkinson Disease Genomics Consortium), additional quality control filtering was applied, including removal of SNPs (i) with MAF < 1%, (ii) displaying an I^2^ heterogeneity value of ≥⍰80 and (iii) where the SNP was not present in at least 9 out of the 13 cohorts included in the meta-analysis.

### Global genetic correlation analysis and estimation of sample overlaps

Across disease trait pairs, LD score regression (LDSC) was used to (i) estimate the observed-scale SNP heritability of each trait (which assumes a continuous liability, and thus may differ from liability-scale estimates of SNP heritability), (ii) determine the global *r*_*g*_ and (iii) estimate sample overlap^66,67^. All disease traits had significant SNP-based heritability (Z-score > 2) and met with the criteria suggested for reliable estimates of genetic correlation, which include: (i) heritability Z-score > 1.5 (optimal > 4), (ii) mean Chi square of test statistics > 1.02, and (iii) intercept estimated from SNP heritability analysis is between 0.9-1.1^68^ (**Supplementary Table 2**). We note that the heritability Z-score of LBD was 2.27, which is below the optimal suggested, and as such, can be expected to produce larger standard errors around estimates of global *r*_*g*_.

Summary statistics for each trait were pre-processed using LDSC’s `munge_sumstats.py` (https://github.com/bulik/ldsc/blob/master/munge_sumstats.py) together with HapMap Project Phase 3 SNPs^69^. For the LD reference panel, 1000 Genomes Project Phase 3 European population SNPs were used^70^. Both HapMap Project Phase 3 SNPs and European LD Scores from the 1000 Genomes Project are made available by the developers of LDSC^66,67^ from the following repository: https://alkesgroup.broadinstitute.org/LDSCORE/ (see **Key resources** for details).

The estimated sample overlap was used as an input for LAVA, given that potential sample overlap between GWASs could impact estimated local *r*_*g*_s^31^. Any shared variance due to sample overlap was modelled as a residual genetic covariance. As performed by Werme *et al*.^31^, a symmetric matrix was constructed, with off-diagonal elements populated by the intercepts for genetic covariance derived from cross-trait LDSC and diagonal elements populated by comparisons of each trait with itself. This symmetric matrix was then converted to a correlation matrix. Importantly, it is not possible to estimate sample overlap with eQTL summary statistics, but given that the cohorts used in the GWASs were different from the cohorts included in the eQTL datasets, we assumed sample overlap between GWASs and eQTL datasets to be negligible. Thus, they were set to 0 in the correlation matrix. However, given the inclusion of GTEx samples in both eQTL datasets and our inability to estimate this overlap, downstream LAVA analyses were performed separately for each eQTL dataset.

### Defining genomic regions for local genetic correlation analysis

#### Between disease traits

Genome-wide significant loci (p < 5 × 10^−8^) were derived from publicly available AD, BIP, LBD, MDD, PD and SCZ GWASs. Genome-wide significant loci were overlapped with linkage disequilibrium (LD) blocks generated by Werme *et al*.^31^ using a genome-wide partitioning algorithm. Briefly, each chromosome was recursively split into blocks using (i) a break point to minimise LD between the resulting blocks and (ii) a minimum size requirement. The resulting LD blocks represent approximately equal-sized, semi-independent blocks of SNPs, with a minimum size requirement of 2,500 SNPs (resulting in an average block size of around 1Mb). Only those LD blocks containing genome-wide significant GWAS loci from at least one trait were carried forward in downstream analyses, resulting in a total of 300 autosomal LD blocks. Of the 22 possible autosomes, 21 contained LD blocks with overlapping loci, with the highest number of LD blocks located in chromosome 1 and 6 (**Supplementary Figure 1**). LD block locations were in reference to build GRCh37 and are presented in the format: LD block identifier, chromosome:start-end.

#### Between disease and gene expression traits

A total of 5 LD blocks, as highlighted by bivariate local *r*_*g*_ analysis of disease traits, were used in this analysis (LD block 681, chr4:90,236,972-91,309,863; LD block 1273, chr8:27,406,512-28,344,176; LD block 1719, chr11:112,755,447-113,889,019; LD block 2281, chr18:52,512,524-53,762,996; LD block 2351, chr19:45,040,933-45,893,307). From these LD blocks of interest, we defined genic regions for all protein-coding, antisense or lincRNA genes that overlapped an LD block of interest. Genic regions were defined as the start and end coordinates of a gene (Ensembl v87, GRCh37) with an additional 100 kb upstream and 100 kb downstream of gene start/end coordinates. We included a 100-kb window as most lead cis-eQTL SNPs (i.e. the SNP with the most significant p-value in a SNP-gene association) lie outside the gene start and end coordinates and are located within 100 kb of the gene (in eQTLGen, 55% of lead-eQTL SNPs were outside the gene body and 92% were within 100 kb from the gene^43^). These genic regions (*n* = 92) were carried forward in downstream analyses. Within these genic regions, eQTL summary statistics were filtered to include all eQTLs (including both significant and non-significant, with significance defined as FDR < 0.05) for the relevant gene (e.g. only SNP-gene pairs that relate to *CLU* in the *CLU* genic region). This was to avoid SNP duplication, which can occur where an eQTL SNP regulates more than one gene.

### Estimating bivariate local genetic correlations

#### Between disease traits

The detection of valid and interpretable local *r*_*g*_ requires the presence of sufficient local genetic signal. For this reason, a univariate test was performed as a filtering step for bivariate local *r*_*g*_ analyses. Bivariate local *r*_*g*_ analyses were only performed for pairs of disease traits which both exhibited a significant univariate local genetic signal (p < 0.05/300, where the denominator represents the total number of tested LD blocks). This step resulted in a total of 1,603 bivariate tests spanning 275 distinct LD blocks. Bivariate results were considered significant when p < 0.05/1603.

#### Between disease and gene expression traits

For each genic region, only those disease traits that were found to have significant local *r*_*g*_ in the associated LD block were carried forward to univariate and bivariate analyses with eQTL summary statistics. As previously described, a univariate test was performed as a filtering step for bivariate local 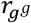 analyses. Thus, bivariate local *r*_*g*_ analyses were only performed (i) if the gene expression trait (i.e. eQTL genes) exhibited a significant univariate local genetic signal and (ii) for pairs of traits (disease and gene expression) which both exhibited a significant univariate local genetic signal. A cut-off of p < 0.05/92 (the denominator represents the total number of tested genic regions) was used to determine univariate significance. A 100-kb window resulted in a total of 354 bivariate tests spanning 55 distinct genic regions. Bivariate results were corrected for multiple testing using two strategies: (i) a more lenient FDR correction and (ii) a more stringent Bonferroni correction (p < 0.05/n_tests, where the denominator represents the total number of bivariate tests). We discuss results passing FDR < 0.05, but we make the results of both correction strategies available (**Supplementary Table 7, Supplementary Table 8**).

We evaluated the effect of window size on bivariate correlations by re-running all analyses using a 50-kb window. Following filtering for significant univariate local genetic signal (as described above), a total of 267 bivariate tests were run spanning 50 distinct genic regions. We detected 110 significant bivariate local *r*_*g*_s (FDR < 0.05), 83 of which were also significant when using a 100-kb window (**Supplementary Figure 6**). We observed strong positive Pearson correlations in local *r*_*g*_ coefficient and p-value estimates across the two window sizes, indicating that our results are robust to the choice of window size (**Supplementary Figure 6**). Of note, p-value estimates between disease and gene expression traits tended to be lower when using the 50-kb window, as compared to the 100-kb window, as evidenced by the fitted line falling below the equivalent of y = x. This observation may be a reflection of stronger *cis*-eQTLs tending to have a smaller distance between SNP and gene^43^. In contrast, p-value estimates between two disease traits were comparable across the two window sizes.

Partial correlations were computed where three-way relationships were observed between 2 disease traits and an eQTL. The partial correlation reflects the correlation between 2 traits (e.g. disease X and Y) that can be explained by a third trait (e.g. eQTL, Z). Thus, a partial correlation approaching 0 suggests that trait Z captures an increasing proportion of the correlation between traits X and Y. Due to the three-way nature of the relationships, 3 possible conformations were possible (i.e. X∼Y|Z, X∼Z|Y and Y∼Z|X); partial correlations were computed for all 3.

### Local multiple regression

For LD blocks with significant bivariate local *r*_*g*_ between one disease trait and ≥ 2 disease traits, multiple regression was used to determine the extent to which the genetic component of the outcome trait could be explained by the genetic components of multiple predictor traits. These analyses permitted exploration of the independent effects of predictor traits on the outcome trait. A predictor trait was considered significant when p < 0.05.

### Sensitivity analysis using by-proxy cases

As UK Biobank (UKBB) by-proxy cases could potentially be mislabelled (i.e. parent of by-proxy case suffered from another type of dementia) and lead to spurious *r*_*g*_s between neurodegenerative traits, we performed replication analyses using GWASs for AD^27^ and PD that excluded UKBB by-proxy cases. LD blocks were filtered to include only those where significant bivariate local *r*_*g*_s were observed between LBD and either by-proxy AD or by-proxy PD GWASs, in addition to between by-proxy AD and by-proxy PD GWASs. These criteria limited the number of LD blocks to 21. Bivariate local correlations were only performed for pairs of traits which both exhibited a significant univariate local genetic signal (p < 0.05/21, where the denominator represents the total number of tested loci), which resulted in a total of 10 bivariate tests spanning 6 distinct loci. We additionally performed multiple regression in LD block 2351 using LBD as the outcome and AD and PD (both excluding UKBB by-proxy cases) as predictors. A predictor trait was considered significant when p < 0.05.

### R packages

All analyses were performed in R (v 4.0.5)^71^. As indicated in the accompanying GitHub repository (https://github.com/RHReynolds/neurodegen-psych-local-corr), all relevant packages were sourced from CRAN, Bioconductor (via BiocManager^72^) or directly from GitHub. Figures were produced using *circlize, ggplot2* and *ggraph*^73–75^. All open-source software used in this paper is listed in **Key resources**.

## Data Availability

All data produced in the present work are either contained in the manuscript or are made available in the accompanying GitHub repository (https://github.com/RHReynolds/neurodegen-psych-local-corr). GWASs used were downloaded from the following sources: Alzheimer's disease, Jansen et al. (https://ctg.cncr.nl/software/summary_statistics); Alzheimer's disease, Kunkle et al. (https://www.niagads.org/igap-rv-summary-stats-kunkle-p-value-data); bipolar disorder (bip2021, https://www.med.unc.edu/pgc/download-results/); Lewy body dementia (https://www.ebi.ac.uk/gwas/studies/GCST90001390); Parkinson's disease (https://pdgenetics.org/resources); major depressive disorder (mdd2019edinburgh, https://www.med.unc.edu/pgc/download-results/), schizophrenia (scz2018clozuk, https://www.med.unc.edu/pgc/download-results/). Cis-eQTL data were obtained from eQTLGen (https://www.eqtlgen.org/cis-eqtls.html) and PsychENCODE (http://resource.psychencode.org). LAVA LD blocks were downloaded from https://github.com/cadeleeuw/lava-partitioning. HapMap Project Phase 3 SNPs and European LD Scores from the 1000 Genomes Project were downloaded from the following Alkes Group repository (https://alkesgroup.broadinstitute.org/LDSCORE/). Human gene annotation (GRCh37, ensembl v87) was downloaded from Ensembl (http://ftp.ensembl.org/pub/grch37/current/gtf/homo_sapiens/).

## Abbreviations

AD: Alzheimer’s disease
BIP: bipolar disorder
bp: base pair
CI: confidence interval
DLB: dementia with Lewy bodies
eQTL: expression quantitative loci
FDR: false discovery rate
GWAS: genome-wide association study
kb: kilobase
LAVA: local analysis of [co]variant annotation
LBD: Lewy body dementia
LD: linkage disequilibrium
LDSC: linkage disequilibrium score regression
MDD: major depressive disorder
PD: Parkinson’s disease
SCZ: schizophrenia
SNP: single nucleotide polymorphism
UKBB: UK Biobank
ρ: rho
*r*_*g*_: genetic correlation

## Code availability

Code used to pre-process GWASs, run genetic correlation analyses and to generate figures for the manuscript are available at: https://github.com/RHReynolds/neurodegen-psych-local-corr (doi:<u>10.5281/zenodo.6587707</u>). All other open-source software used in this paper is listed in **Key resources**.

## Data availability

Analyses in this study relied on publicly available data, all of which are listed in **Key resources**. In the case of the PD GWAS without UK Biobank (UKBB) data, summary statistics were kindly provided by the International Parkinson Disease Genomics Consortium: https://pdgenetics.org/.

## Key resources

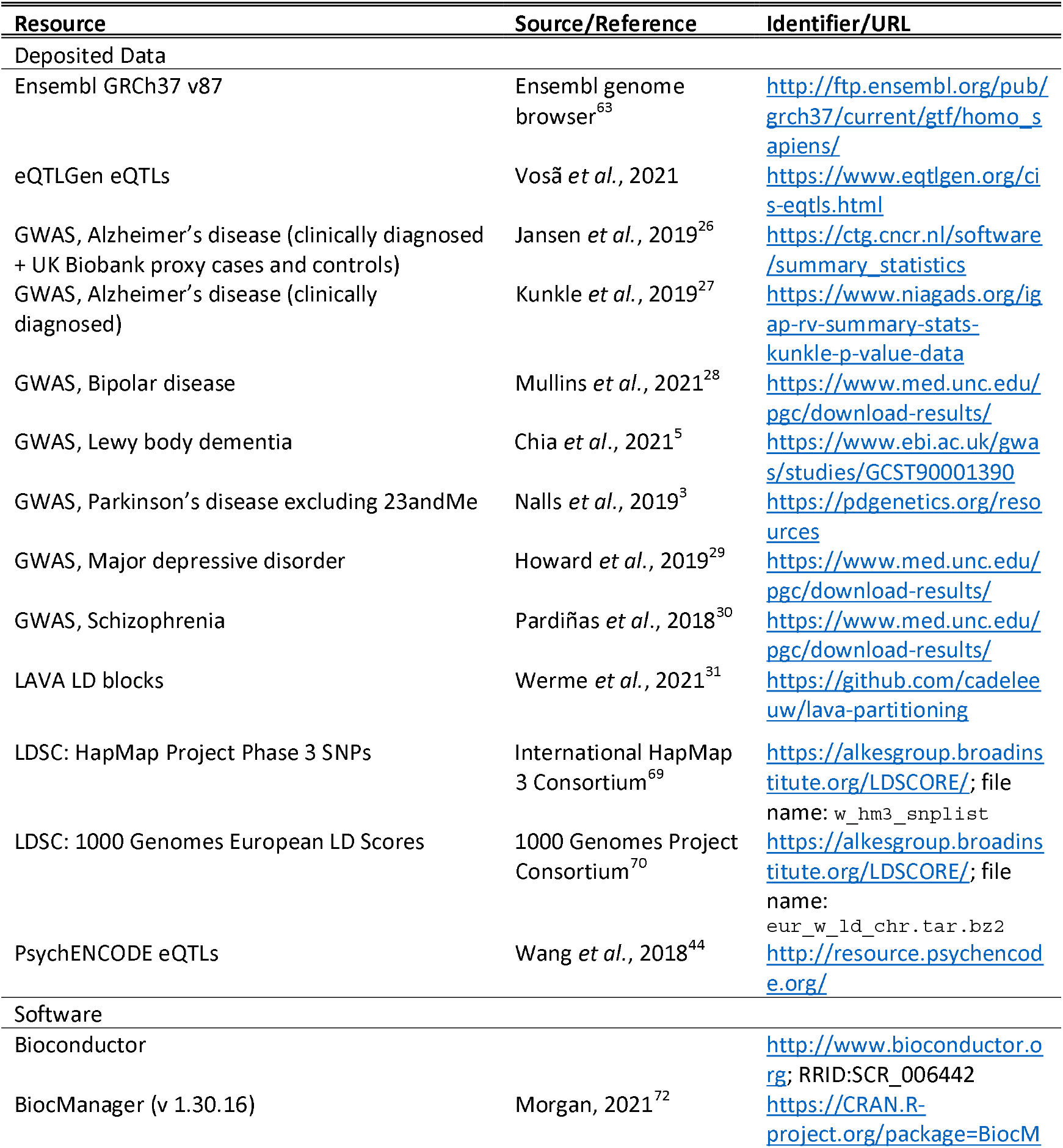
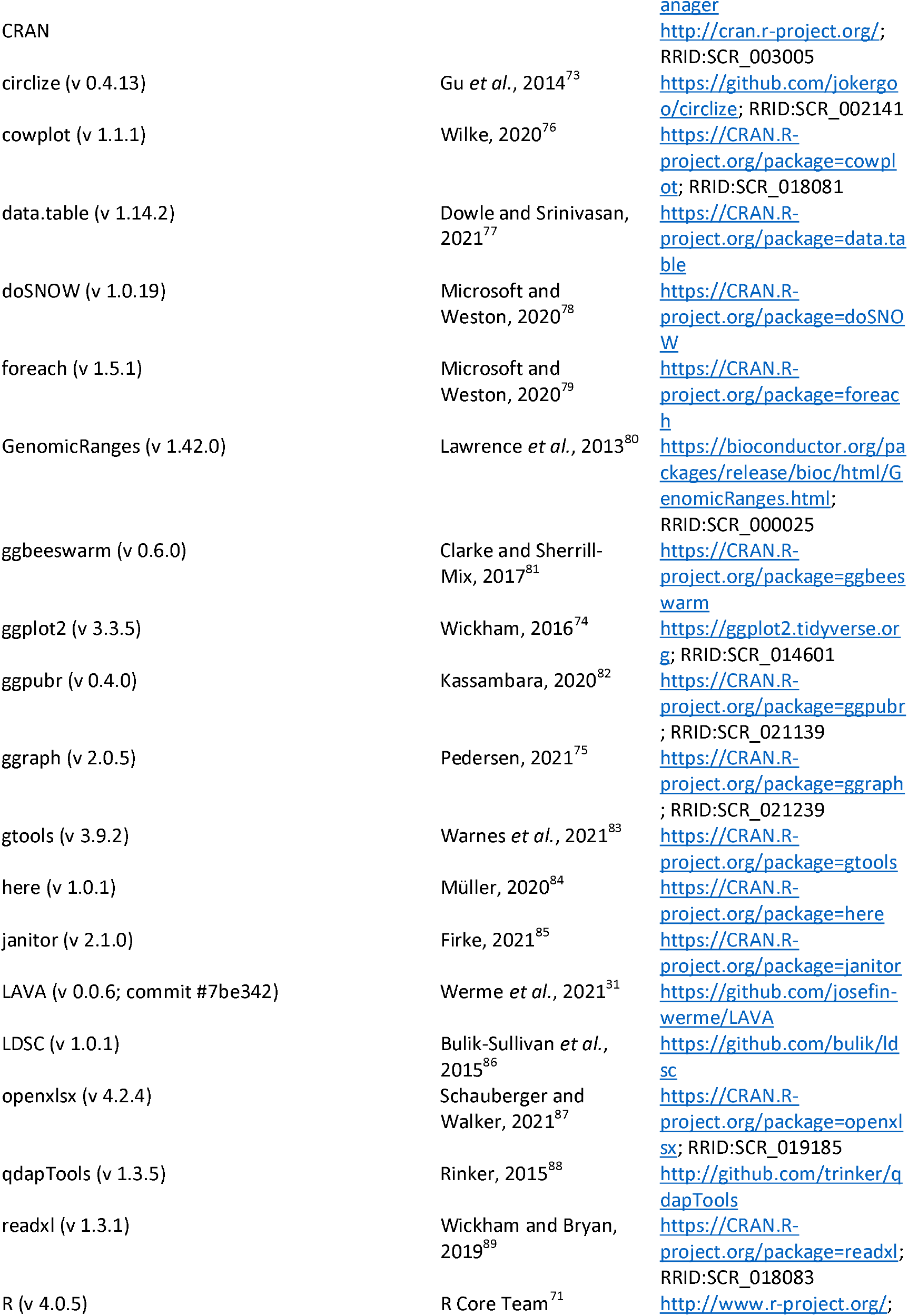
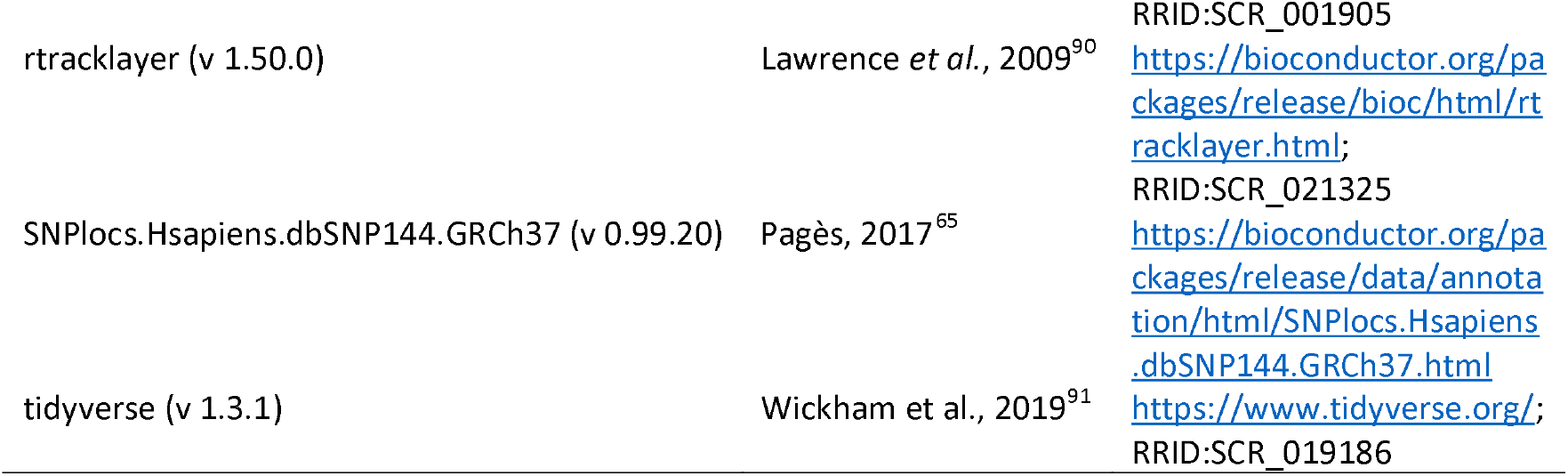

## Acknowledgements

We are grateful to Dr Cornelis Blauwendraat for feedback throughout this project. We would also like to thank the scientific community behind all the GWAS datasets, as well as data scientists developing the above-mentioned analysis tools, for making them publicly available and thus enabling the completion of this study.

## Author contributions

RHR, SAGT and MR conceived and designed the study. RHR and AZW analysed data and drafted the figures. FLD, MS and SAGT consulted on the statistical analysis. SWS and MR provided clinical insight to data interpretation. RHR wrote the initial manuscript. All authors contributed to the critical analysis and revision of the manuscript.

## Supplementary Figures

**Supplementary Figure 1.**
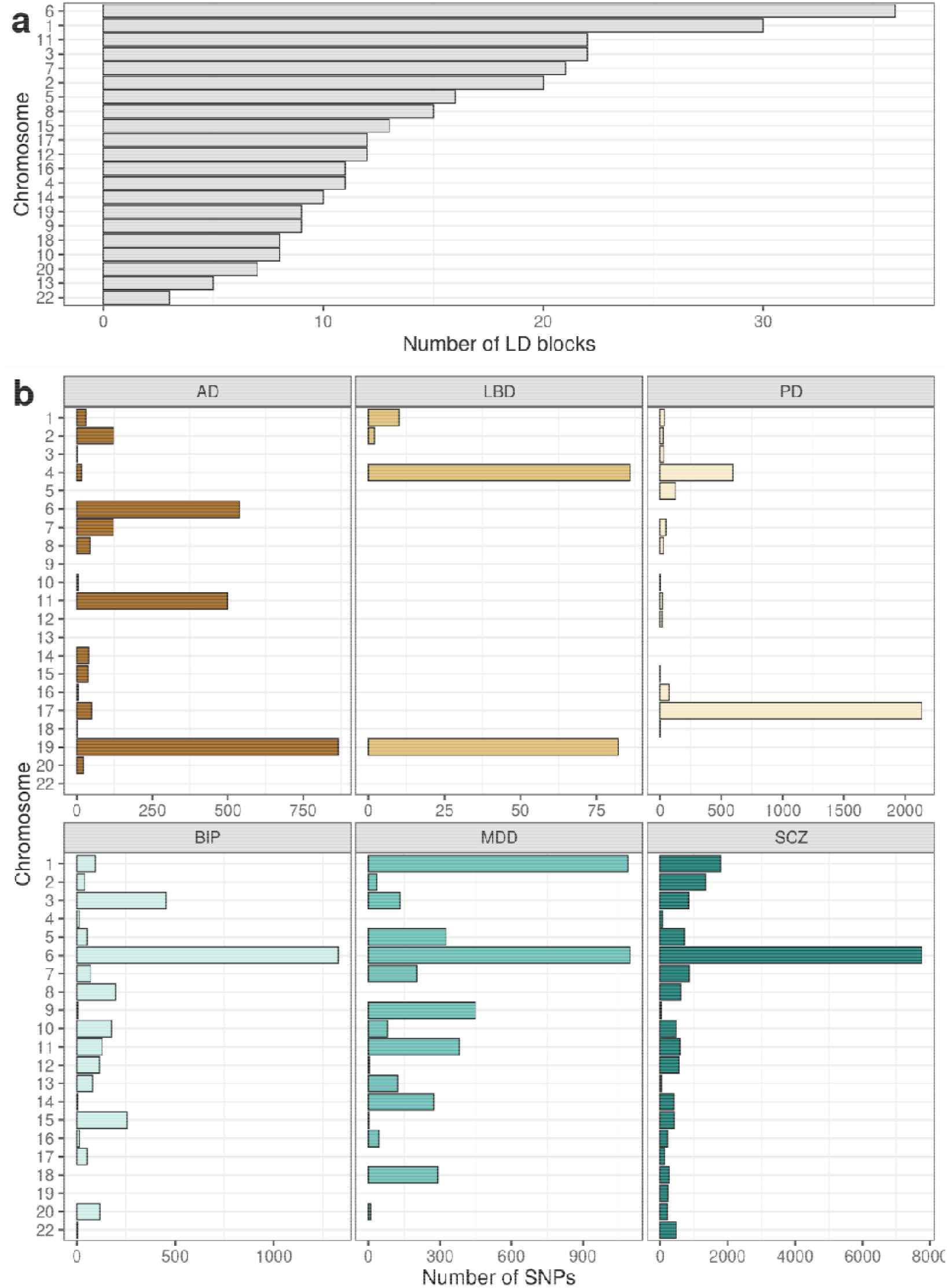
**(a)** Number of LD blocks containing genome-wide significant loci per chromosome. Chromosomes have been ordered by the total number of LD blocks in each chromosome. **(b)** Number of genome-wide significant AD, BIP, LBD, MDD, PD and SCZ SNPs per autosome.

**Supplementary Figure 2.**
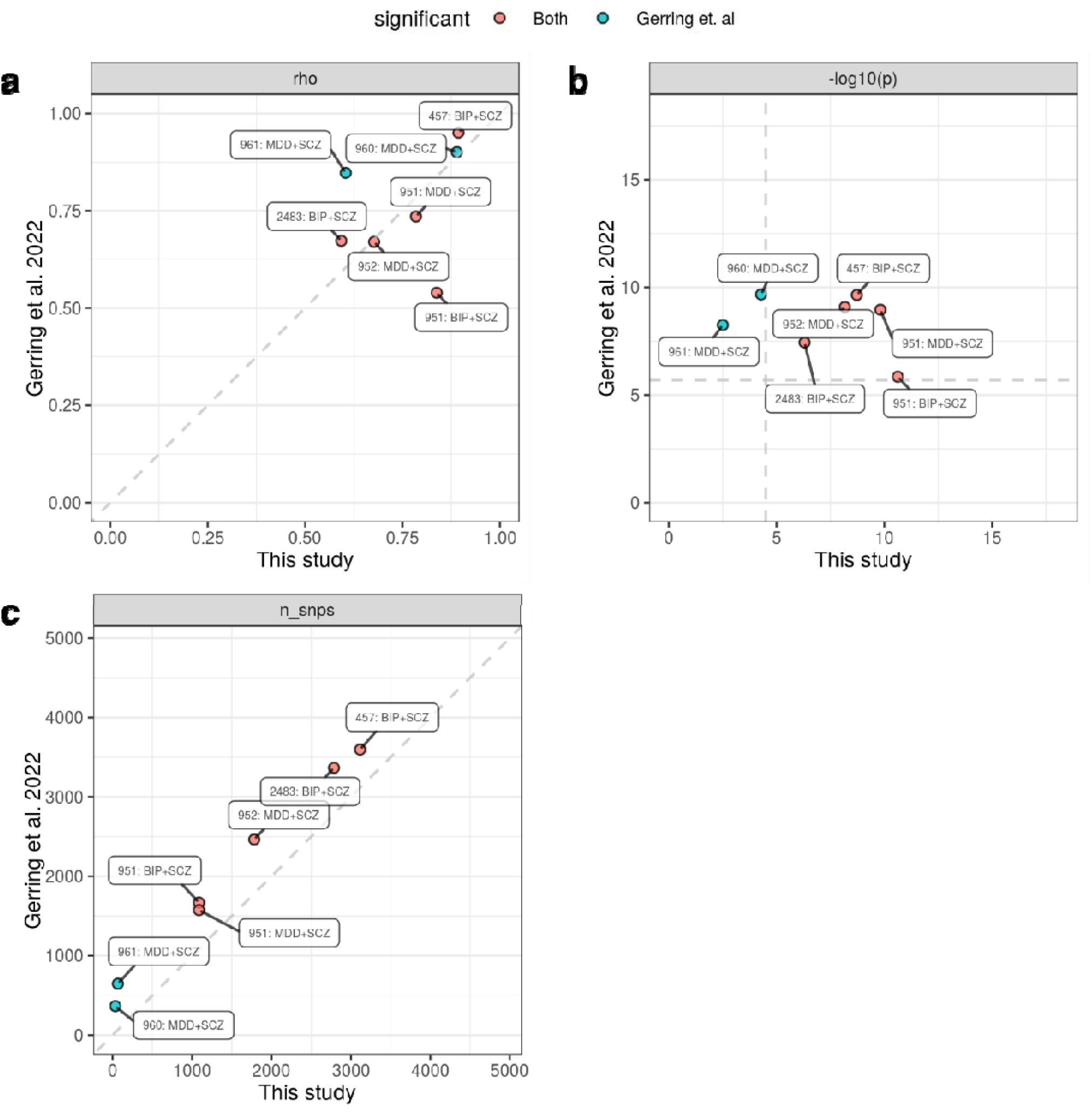
Scatter plot of **(a)** the standardised coefficient for (rho, ρ), **(b)** -log10(p-value) and **(c)** number of overlapping SNPs for each pair of traits that overlapped between our study and the study by Gerring *et al*.^18^ In **(a)** the dashed lines indicate significance thresholds in each study (Gerring *et al*., p < 0.05/24,054; this study, p < 0.05/1603) and in **(b, c)** the dashed line represents the line y = x. Points in both plots are coloured by whether they passed significance thresholds in both studies or only the Gerring *et al*. study. Points are labelled by LD block and trait pair. Coordinates for LD blocks (in the format chromosome:start-end, GRCh37): 457, chr3:36,840,137-38,729,767; 951, chr6:26,396,201-27,261,035; 952, chr6:27,261,036-28,666,364; 960, chr6: 31,320,269-31,427,209; 961, chr6:31,427,210-32,208,901; 2483, chr22:38,718,590-40,378,783.

**Supplementary Figure 3.**
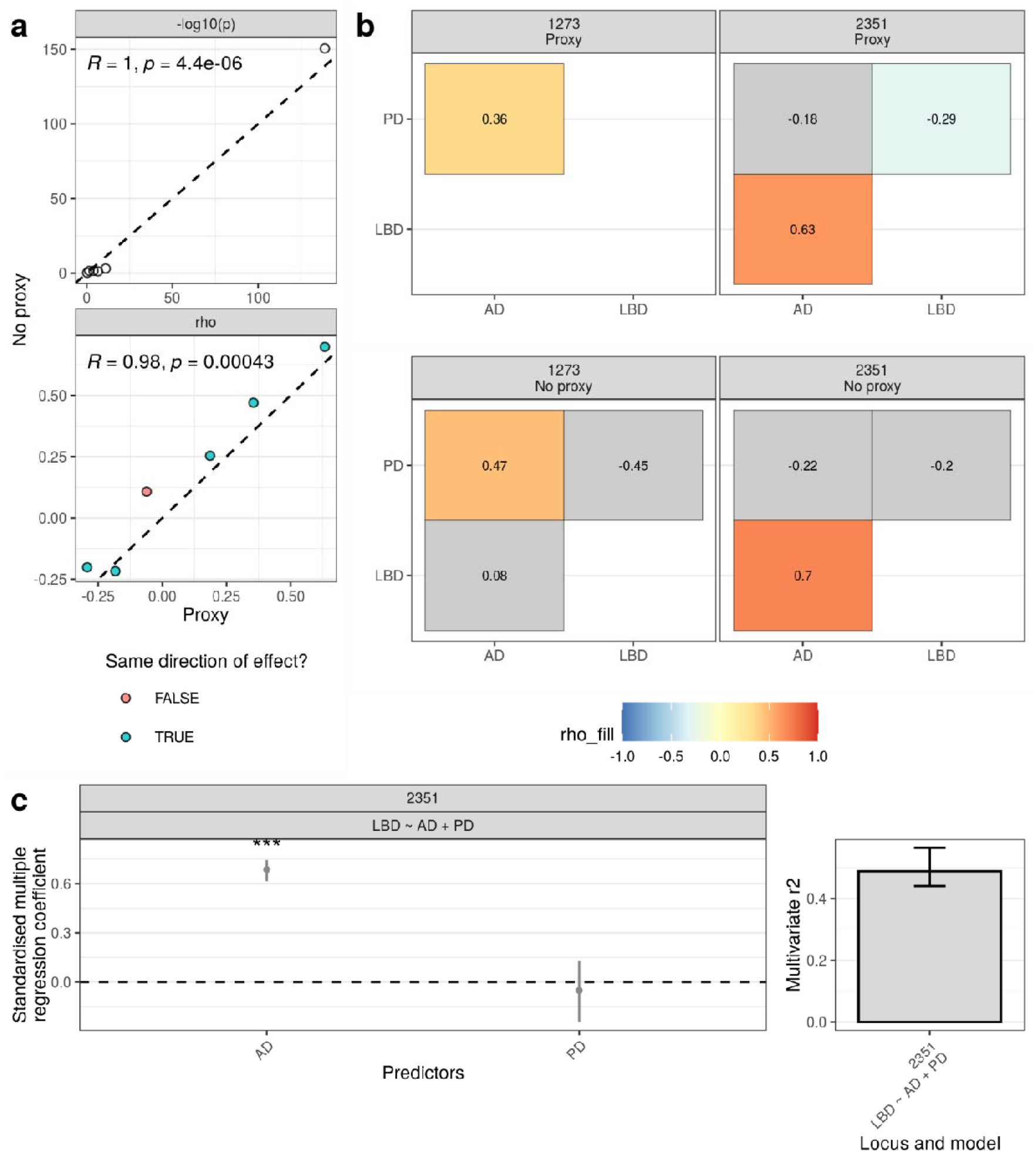
Impact of excluding UK Biobank by-proxy cases on local genetic correlations and multiple regression. **(a)**Scatter plot of -log10(p-value) and the standardised coefficient for (rho, ρ) for each pair of traits with sufficient univariate signal to carry out a bivariate test using AD/PD GWASs with or without by-proxy cases. In each panel, Pearson’s coefficient (R) and associated p-value (p) are displayed. The black dashed line represents the line y = x. Points are coloured, where applicable, by whether they share the same direction of effect. **(b)** Significant bivariate local genetic correlations using AD/PD GWASs with or without by-proxy cases (as indicated in panel headers). Heatmaps show the rho for all tested associations within the LD block, with significant negative and positive correlations indicated by blue and red fill, respectively. Non-significant correlations have a grey fill. **(c)** Results of multiple regression model across LD block 2351. Plot (left) of standardised coefficients for each predictor in multiple regression model in LD block 2351, with whiskers spanning the 95% confidence interval for the coefficients. Plot (right) of multivariate for LD block 2351, where multivariate represents the proportion of variance in genetic signal for LBD explained by AD and PD simultaneously. Whiskers span the 95% confidence interval for the multivariate. ***, p < 0.001. Coordinates for LD blocks (in the format chromosome:start-end, GRCh37): 1273, chr8:27,406,512-28,344,176; 2351, chr19:45,040,933-45,893,307.

**Supplementary Figure 4.**
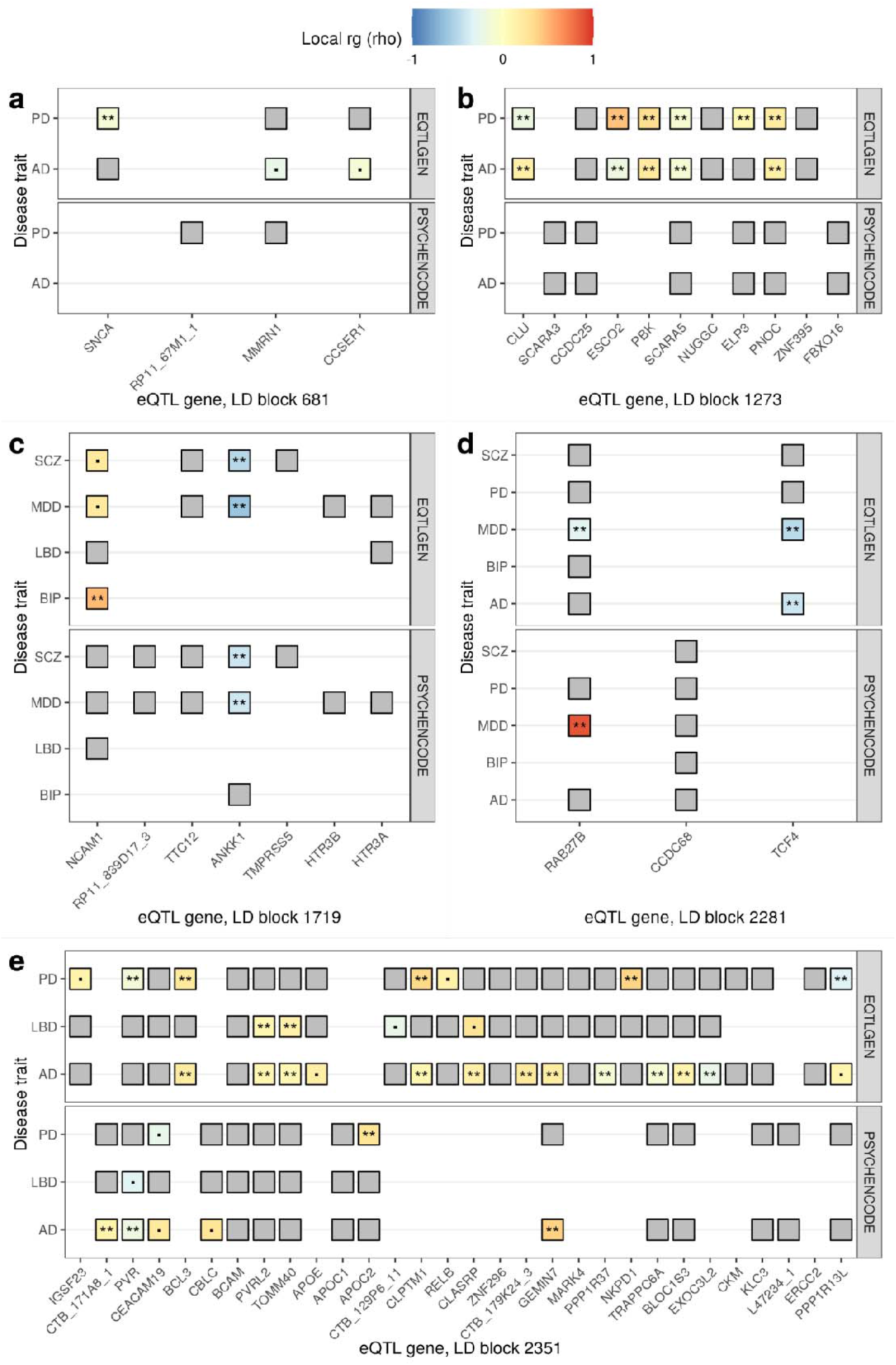
Local gene expression and disease trait correlations across 5 LD blocks of interest. Heatmaps of the standardised coefficient for *r*_*g*_ (rho) for all tested gene expression-disease trait correlation within LD block **(a)** 681, **(b)** 1273, **(c)** 1719, **(d)** 2281 and **(e)** 2351. All negative and positive *r*_*g*_s with p < 0.05 are indicated by blue and red colour, respectively, while the remainder have a grey fill. Significant local *r*_*g*_ s (FDR < 0.05) are indicated by two asterisks (**), while nominally significant local *r*_*g*_s (p < 0.05) are indicated with a black square (⍰). Genes are ordered left to right on the x-axis by the genomic coordinate of their gene start. Coordinates for LD blocks (in the format chromosome:start-end, GRCh37): 681, chr4:90,236,972-91,309,863; 1273, chr8:27,406,512-28,344,176; 1719, chr11:112,755,447-113,889,019; 2281, chr18:52,512,524-53,762,996; 2351, chr19:45,040,933-45,893,307.

**Supplementary Figure 5.**
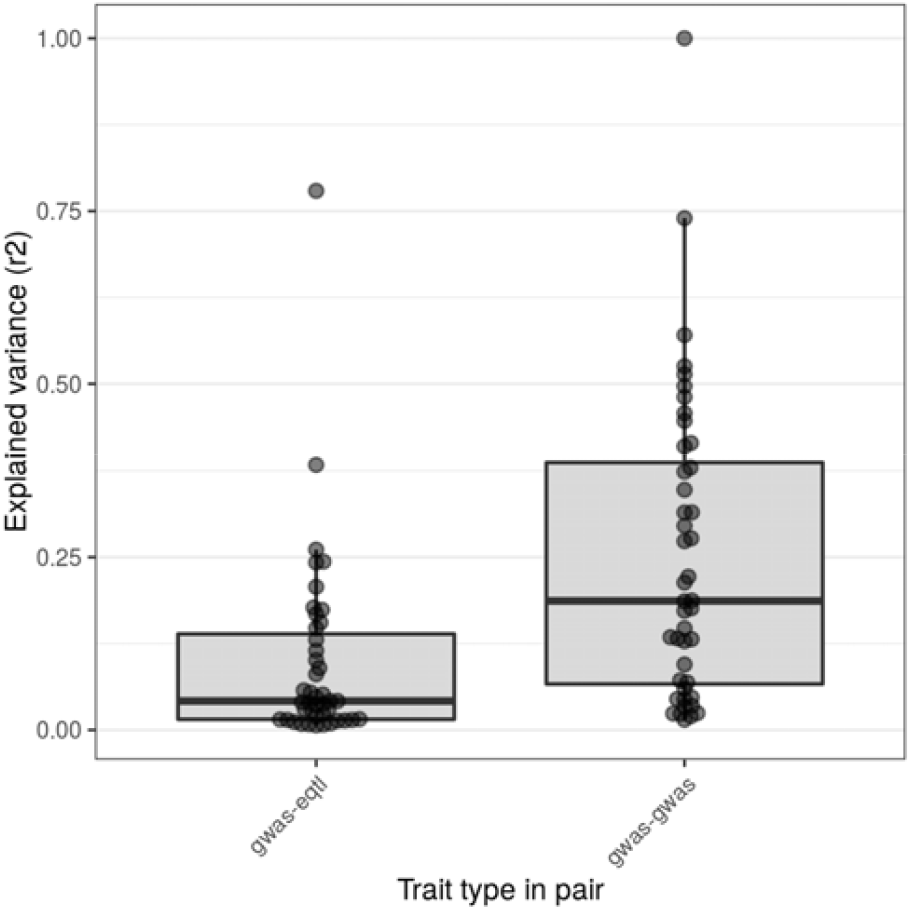
Explained variance in trait pairs with different trait types. Boxplot of explained variance (*r*^2^, the proportion of variance in genetic signal of one disease trait in a pair explained by the other) in trait pairs involving a disease and gene expression trait (gwas-eqtl) or two disease traits (gwas-gwas). Only local *r*_*g*_s that passed significance are plotted (FDR < 0.05; N, local *r*_*g*_s = 87).

**Supplementary Figure 6.**
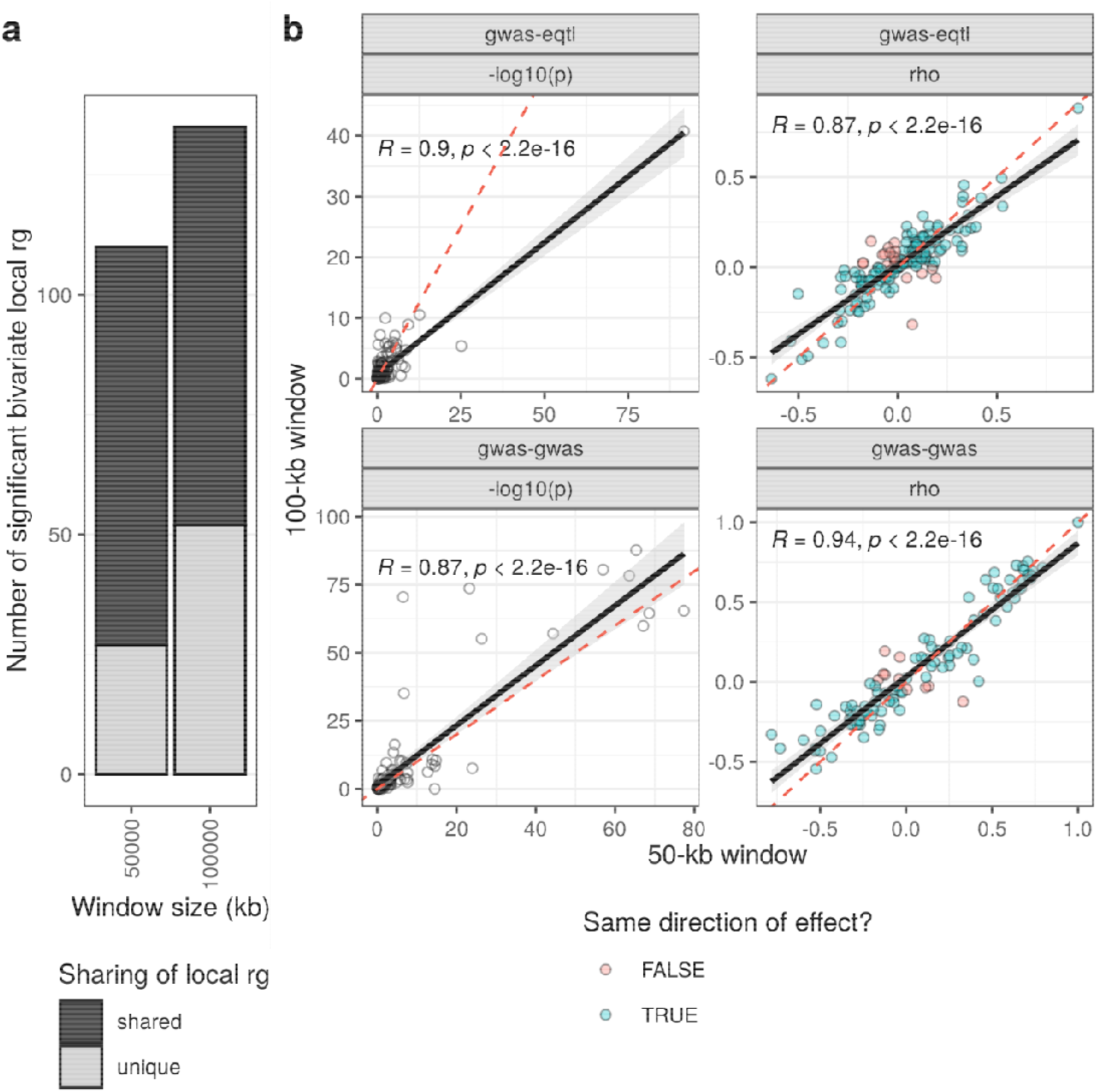
Effect of window size on local genetic correlations. (a) Number of significant bivariate local ’s across window sizes. Bars are coloured by whether ’s are significant across both window sizes (shared) or only one (unique). (b) Scatter plot of -log10(p-value) and the standardised coefficient for (rho, ρ) for each pair of traitss that could be tested across genic regions with a 50-kb or 100-kb window. Panels indicate whether the pair of traits included a disease and gene expression trait (gwas-eqtl) or two disease traits (gwas-gwas). Points are coloured by whether they share the same direction of effect. The black line represents a linear model fitted to the data, with the 99% confidence interval indicated with a grey fill. Further, Pearson’s coefficient (R) and associated p-value (p) are displayed. The red dashed line represents the line y = x.

## Supplementary Tables

**Supplementary Table 1** LD blocks, their associated disease traits (as determined by overlap of genome-wide significant SNPs) and overlapping genes.

**Supplementary Table 2** Results of LDSC using the six disease traits.

**Supplementary Table 3** Results of LAVA using the six disease traits.

**Supplementary Table 4** Overlaps between results of LAVA and results from Drange *et al*^37^, Smeland *et al*^*24*^. and Gerring *et al*.^18^

**Supplementary Table 5** Results of LAVA using GWASs for AD and PD that exclude UK Biobank by-proxy cases.

**Supplementary Table 6** Results of multiple regression analyses.

**Supplementary Table 7** Results of LAVA using disease and gene expression traits (100-kb window). Sheets containing bivariate results for each LD block also contain (a) locus plot of genic regions (including 100-kb window). Significant bivariate local genetic correlations between a disease and gene expression trait are highlighted in blue (FDR < 0.05). (b) Edge diagrams for genic regions where a significant bivariate local genetic correlation was observed between a disease and gene expression trait (FDR < 0.05). Edges display the standardised coefficient for genetic correlation (rho) for significant bivariate local genetic correlations, with negative and positive correlations indicated by blue and red colour, respectively. GWAS and eQTL nodes are indicated by grey and white fill, respectively.

**Supplementary Table 8** Results of LAVA using disease and gene expression traits (50-kb window). Sheets containing bivariate results for each LD block also contain (a) locus plot of genic regions (including 50-kb window). Significant bivariate local genetic correlations between a disease and gene expression trait are highlighted in blue (FDR < 0.05). (b) Edge diagrams for genic regions where a significant bivariate local genetic correlation was observed between a disease and gene expression trait (FDR < 0.05). Edges display the standardised coefficient for genetic correlation (rho) for significant bivariate local genetic correlations, with negative and positive correlations indicated by blue and red colour, respectively. GWAS and eQTL nodes are indicated by grey and white fill, respectively.

**Supplementary Table 9** Results of partial correlations using disease and gene expression traits (100-kb window).

## Supplementary Note

### Comparison of local *r*_*g*_s to existing reports of local genetic relations

#### Methods

We compared local *r*_*g*_s to existing results from studies of: (i) AD and PD using rho-HESS^19^; (ii) AD and BIP^21^, AD and MDD^23^, PD and SCZ^24^, all of which used a conditional/conjunctional FDR approach (conditional FDR is an extension of the standard FDR method, and re-ranks the test statistics of a primary phenotypes based on the strength of the association with a secondary phenotype, while conjunctional FDR is used post-hoc to identify shared genetic loci); and (iii) 10 psychiatric disorders and 10 substance abuse phenotypes using LAVA^18^. For all comparisons, genomic coordinates were used to overlap either SNPs^21,24^ or genomic regions^18,19^ with LD blocks. SNPs were converted from rsIDs to their GRCh37 genomic coordinates using the SNPlocs.Hsapiens.dbSNP144.GRCh37 package^65^. In all comparisons, local *r*_*g*_s from this study were filtered to include only relevant disease traits.

#### Results

We were unable to replicate the genetic overlaps between AD and PD reported in the *HLA* (specifically in chr6: 31,571,218-32,682,664^19^) and *MAPT* (rs393152 shown to increase risk of both AD and PD^20^) loci. In the *MAPT* locus, this was due to insufficient univariate signal for AD in the LD block containing *MAPT* (LD block 2207, chr17: 43,460,501-44,865,832), while in the *HLA* locus, several of the overlapping LD blocks (LD block 961-6, ranging across chr6: 31,427,210-32,682,213) had too few overlapping SNPs between the 6 disease traits.

We were unable to replicate any of the associations reported using a conditional/conjunctional FDR approach. For 4 associations (rs11649476 in AD and BIP^21^; rs5011436 in AD and MDD^23^; rs2979160 and rs4921739 in PD and SCZ^24^) this was due to a lack of overlap with the 300 LD blocks tested. For 1 association (rs62333164 in PD and SCZ^24^), this was due to insufficient univariate signal for SCZ in the overlapping LD block (LD block 752, chr4:169555115-170682809). Of the 9 loci that were jointly associated with PD and SCZ^24^, 6 intersected with LD blocks where we performed a bivariate test for PD and SCZ, but none of the bivariate tests passed the threshold for significance (p < 0.05/1,603**; Supplementary Table 4, Supplementary Table 5**).

Of the 14 LD blocks found to contain significant local *r*_*g*_s between any of BIP, MDD and SCZ in the study by Gerring *et al*., 6 LD blocks intersected with LD blocks where we performed a bivariate test using the same phenotype pairs. We were able to replicate 5 of the 7 overlapping local *r*_*g*_s (BIP and SCZ in LD block 457; SCZ and BIP or MDD in LD block 951; MDD and SCZ in LD block 952; and BIP and SCZ in LD block 2483; **Supplementary Figure 2; Supplementary Table 4**). Our inability to replicate 2 local s could be due to: (i) the lower number of overlapping SNPs present in the tested LD blocks in our study compared to Gerring *et al*. (**Supplementary Figure 2c**); and/or (ii) differences in GWASs used (while both studies used the same MDD and SCZ GWASs, our study did not include 23andMe data from the MDD GWAS and used a more recent BIP GWAS).

## References

1. Robinson, J. L. et al. Neurodegenerative disease concomitant proteinopathies are prevalent, age-related and APOE4-associated. Brain J. Neurol. 141, 2181–2193 (2018).

2. De Jager, P. L., Yang, H.-S. & Bennett, D. A. Deconstructing and targeting the genomic architecture of human neurodegeneration. Nat. Neurosci. 21, 1310–1317 (2018).

3. Nalls, M. A. et al. Identification of novel risk loci, causal insights, and heritable risk for Parkinson’s disease: a meta-analysis of genome-wide association studies. Lancet Neurol. 18, 1091–1102 (2019).

4. Guerreiro, R. et al. Investigating the genetic architecture of dementia with Lewy bodies: a two-stage genome-wide association study. Lancet Neurol. 17, 64–74 (2018).

5. Chia, R. et al. Genome sequencing analysis identifies new loci associated with Lewy body dementia and provides insights into its genetic architecture. Nat. Genet. 53, 294–303 (2021).

6. Masters, C. L. et al. Alzheimer’s disease. Nat. Rev. Dis. Primer 1, 15056 (2015).

7. Poewe, W. et al. Parkinson disease. Nat. Rev. Dis. Primer 3, 17013 (2017).

8. Kuring, J. K., Mathias, J. L. & Ward, L. Prevalence of Depression, Anxiety and PTSD in People with Dementia: a Systematic Review and Meta-Analysis. Neuropsychol. Rev. 28, 393–416 (2018).

9. Reijnders, J. S. A. M., Ehrt, U., Weber, W. E. J., Aarsland, D. & Leentjens, A. F. G. A systematic review of prevalence studies of depression in Parkinson’s disease. Mov. Disord. Off. J. Mov. Disord. Soc. 23, 183–9; quiz 313 (2008).

10. Weintraub, D. et al. The neuropsychiatry of Parkinson’s disease: advances and challenges. Lancet Neurol. 21, 89–102 (2022).

11. Ribe, A. R. et al. Long-term risk of dementia in persons with schizophrenia: A danish population-based cohort study. JAMA Psychiatry 72, 1095–1101 (2015).

12. Stroup, T. S. et al. Age-Specific Prevalence and Incidence of Dementia Diagnoses among Older US Adults with Schizophrenia. JAMA Psychiatry 78, 632–641 (2021).

13. Gustafsson, H., Nordström, A. & Nordström, P. Depression and subsequent risk of Parkinson disease A nationwide cohort study. Neurology 84, 2422–2429 (2015).

14. Kazmi, H. et al. Late onset depression: Dopaminergic deficit and clinical features of prodromal Parkinson’s disease: A cross-sectional study. J. Neurol. Neurosurg. Psychiatry 92, 158–164 (2021).

15. Bellou, E., Stevenson-Hoare, J. & Escott-Price, V. Polygenic risk and pleiotropy in neurodegenerative diseases. Neurobiol. Dis. 142, 104953 (2020).

16. Brainstorm Consortium et al. Analysis of shared heritability in common disorders of the brain. Science 360, eaap8757 (2018).

17. van Rheenen, W., Peyrot, W. J., Schork, A. J., Lee, S. H. & Wray, N. R. Genetic correlations of polygenic disease traits: from theory to practice. Nat. Rev. Genet. 20, 567–581 (2019).

18. Gerring, Z. F., Thorp, J. G., Gamazon, E. R. & Derks, E. M. A Local Genetic Correlation Analysis Provides Biological Insights Into the Shared Genetic Architecture of Psychiatric and Substance Use Phenotypes. Biol. Psychiatry 92, 583–591 (2022).

19. Stolp Andersen, M., Tan, M., Holtman, I. R., Hardy, J. & Pihlstrøm, L. Dissecting the limited genetic overlap of Parkinson’s and Alzheimer’s disease. Ann. Clin. Transl. Neurol. 1–7 (2022) doi:10.1002/acn3.51606.

20. Desikan, R. S. et al. Genetic overlap between Alzheimer’s disease and Parkinson’s disease at the MAPT locus. Mol. Psychiatry 20, 1588–1595 (2015).

21. Drange, O. K. et al. Genetic overlap between alzheimer’s disease and bipolar disorder implicates the MARK2 and VAC14 genes. Front. Neurosci. 13, 1–11 (2019).

22. Lutz, M. W., Sprague, D., Barrera, J. & Chiba-Falek, O. Shared genetic etiology underlying Alzheimer’s disease and major depressive disorder. Transl. Psychiatry 10, (2020).

23. Monereo-Sánchez, J. et al. Genetic Overlap Between Alzheimer’s Disease and Depression Mapped Onto the Brain. Front. Neurosci. 15, 653130 (2021).

24. Smeland, O. B. et al. Genome-wide Association Analysis of Parkinson’s Disease and Schizophrenia Reveals Shared Genetic Architecture and Identifies Novel Risk Loci. Biol. Psychiatry 89, 227–235 (2021).

25. GBD 2019 Diseases and Injuries Collaborators. Global burden of 369 diseases and injuries in 204 countries and territories, 1990-2019: a systematic analysis for the Global Burden of Disease Study 2019. Lancet Lond. Engl. 396, 1204–1222 (2020).

26. Jansen, I. E. et al. Genome-wide meta-analysis identifies new loci and functional pathways influencing Alzheimer’s disease risk. Nat. Genet. 51, 404–413 (2019).

27. Kunkle, B. W. et al. Genetic meta-analysis of diagnosed Alzheimer’s disease identifies new risk loci and implicates Aβ, tau, immunity and lipid processing. Nat. Genet. 51, 414–430 (2019).

28. Mullins, N. et al. Genome-wide association study of more than 40,000 bipolar disorder cases provides new insights into the underlying biology. Nat. Genet. 53, 817–829 (2021).

29. Howard, D. M. et al. Genome-wide meta-analysis of depression identifies 102 independent variants and highlights the importance of the prefrontal brain regions. Nat. Neurosci. 22, 343–352 (2019).

30. Pardiñas, A. F. et al. Common schizophrenia alleles are enriched in mutation-intolerant genes and in regions under strong background selection. Nat. Genet. 50, 381–389 (2018).

31. Werme, J., van der Sluis, S., Posthuma, D. & de Leeuw, C. A. An integrated framework for local genetic correlation analysis. Nat. Genet. 54, 274–282 (2022).

32. Shi, H., Mancuso, N., Spendlove, S. & Pasaniuc, B. Local Genetic Correlation Gives Insights into the Shared Genetic Architecture of Complex Traits. Am. J. Hum. Genet. 101, 737–751 (2017).

33. Zhang, Y. et al. SUPERGNOVA: local genetic correlation analysis reveals heterogeneous etiologic sharing of complex traits. Genome Biol. 22, 262 (2021).

34. Jellinger, K. A. Dementia with Lewy bodies and Parkinson’s disease-dementia: current concepts and controversies. J. Neural Transm. 125, 615–650 (2018).

35. Poewe, W. et al. Parkinson disease. Nat. Rev. Dis. Primer 3, 17013 (2017).

36. Erkkinen, M. G., Kim, M. & Geschwind, M. D. Clinical Neurology and Epidemiology of the Major Neurodegenerative Diseases. Cold Spring Harb. Perspect. Biol. 10, (2018).

37. Drange, O. K. et al. Genetic overlap between alzheimer’s disease and bipolar disorder implicates the MARK2 and VAC14 genes. Front. Neurosci. 13, 1–11 (2019).

38. Urs, N. M., Peterson, S. M. & Caron, M. G. New Concepts in Dopamine D2Receptor Biased Signaling and Implications for Schizophrenia Therapy. Biol. Psychiatry 81, 78–85 (2017).

39. Underwood, R. et al. The GTPase Rab27b regulates the release, autophagic clearance, and toxicity of α-synuclein. J. Biol. Chem. 295, 8005–8016 (2020).

40. Foster, E. M., Dangla-Valls, A., Lovestone, S., Ribe, E. M. & Buckley, N. J. Clusterin in Alzheimer’s Disease: Mechanisms, Genetics, and Lessons From Other Pathologies. Front. Neurosci. 13, 164 (2019).

41. Bertram, L., McQueen, M. B., Mullin, K., Blacker, D. & Tanzi, R. E. Systematic meta-analyses of Alzheimer disease genetic association studies: The AlzGene database. Nat. Genet. 39, 17–23 (2007).

42. Singh, T. et al. Rare loss-of-function variants in SETD1A are associated with schizophrenia and developmental disorders. Nat. Neurosci. 19, 571–577 (2016).

43. Võsa, U. et al. Large-scale cis- and trans-eQTL analyses identify thousands of genetic loci and polygenic scores that regulate blood gene expression. Nat. Genet. 53, 1300–1310 (2021).

44. Wang, D. et al. Comprehensive functional genomic resource and integrative model for the human brain. Science 362, (2018).

45. Leeuw, C. de, Werme, J., Savage, J. E., Peyrot, W. J. & Posthuma, D. Reconsidering the validity of transcriptome-wide association studies. 2021.08.15.456414 Preprint at https://doi.org/10.1101/2021.08.15.456414 (2022).

46. Yao, D. W., O’Connor, L. J., Price, A. L. & Gusev, A. Quantifying genetic effects on disease mediated by assayed gene expression levels. Nat. Genet. 52, 626–633 (2020).

47. Ponce, G. et al. The ANKK1 kinase gene and psychiatric disorders. Neurotox. Res. 16, 50–59 (2009).

48. Li, J. Y. et al. Scara5 Is a Ferritin Receptor Mediating Non-Transferrin Iron Delivery. Dev. Cell 16, 35–46 (2009).

49. Crielaard, B. J., Lammers, T. & Rivella, S. Targeting iron metabolism in drug discovery and delivery. Nat. Rev. Drug Discov. 16, 400–423 (2017).

50. Han, Z., Li, L., Huang, Y., Zhao, H. & Luo, Y. PBK/TOPK: A Therapeutic Target Worthy of Attention. Cells 10, 371 (2021).

51. Dougherty, J. D. et al. PBK/TOPK, a Proliferating Neural Progenitor-Specific Mitogen-Activated Protein Kinase Kinase. J. Neurosci. 25, 10773–10785 (2005).

52. Nho, K. et al. Association analysis of rare variants near the APOE region with CSF and neuroimaging biomarkers of Alzheimer’s disease. BMC Med. Genomics 10, (2017).

53. Zhou, X. et al. Non-coding variability at the APOE locus contributes to the Alzheimer’s risk. Nat. Commun. 10, 1–16 (2019).

54. Locascio, J. J. et al. Association between α-synuclein blood transcripts and early, neuroimaging-supported Parkinson’s disease. Brain 138, 2659–2671 (2015).

55. Guerreiro, R. et al. Genome-wide analysis of genetic correlation in dementia with Lewy bodies, Parkinson’s and Alzheimer’s diseases. Neurobiol. Aging 38, 214.e7-214.e10 (2016).

56. Moskvina, V. et al. Analysis of genome-wide association studies of alzheimer disease and of parkinson disease to determine if these 2 diseases share a common genetic risk. JAMA Neurol. 70, 1268–1276 (2013).

57. Gibson, J. et al. Assessing the presence of shared genetic architecture between Alzheimer’s disease and major depressive disorder using genome-wide association data. Transl. Psychiatry 7, (2017).

58. McKeith, I. G. et al. Diagnosis and management of dementia with Lewy bodies: Fourth consensus report of the DLB Consortium. Neurology 89, 88–100 (2017).

59. Ozaki, Y. et al. DRD2 methylation to differentiate dementia with Lewy bodies from Parkinson’s disease. Acta Neurol. Scand. 141, 177–182 (2020).

60. Piggott, M. A. et al. Selective loss of dopamine D2 receptors in temporal cortex in dementia with Lewy bodies, association with cognitive decline. Synapse 61, 903–911 (2007).

61. Foo, J. N. et al. Identification of Risk Loci for Parkinson Disease in Asians and Comparison of Risk Between Asians and Europeans: A Genome-Wide Association Study. JAMA Neurol. 77, 746–754 (2020).

62. Brown, B. C., Asian Genetic Epidemiology Network Type 2 Diabetes Consortium, Ye, C. J., Price, A. L. & Zaitlen, N. Transethnic Genetic-Correlation Estimates from Summary Statistics. Am. J. Hum. Genet. 99, 76–88 (2016).

63. Zerbino, D. R. et al. Ensembl 2018. Nucleic Acids Res. 46, D754–D761 (2018).

64. Broekema, R. V., Bakker, O. B. & Jonkers, I. H. A practical view of fine-mapping and gene prioritization in the post-genome-wide association era. Open Biol. 10, 190221.

65. Pagès, H. SNPlocs.Hsapiens.dbSNP144.GRCh37: SNP locations for Homo sapiens (dbSNP Build 144). R Package Version 09920 (2017).

66. Bulik-Sullivan, B. et al. LD score regression distinguishes confounding from polygenicity in genome-wide association studies. Nat. Genet. 47, 291–295 (2015).

67. Bulik-Sullivan, B. et al. An atlas of genetic correlations across human diseases and traits. Nat. Genet. 47, 1236–1241 (2015).

68. Zheng, J. et al. LD Hub: a centralized database and web interface to perform LD score regression that maximizes the potential of summary level GWAS data for SNP heritability and genetic correlation analysis. Bioinforma. Oxf. Engl. 33, 272–279 (2017).

69. International HapMap 3 Consortium et al. Integrating common and rare genetic variation in diverse human populations. Nature 467, 52–8 (2010).

70. 1000 Genomes Project Consortium et al. A global reference for human genetic variation. Nature 526, 68–74 (2015).

71. R Core Team. R: A language and environment for statistical computing. Preprint at (2021).

72. Morgan, M. BiocManager: Access the Bioconductor Project Package Repository. Preprint at (2021).

73. Gu, Z., Gu, L., Eils, R., Schlesner, M. & Brors, B. circlize Implements and enhances circular visualization in R. Bioinforma. Oxf. Engl. 30, 2811–2 (2014).

74. Wickham, H. ggplot2: Elegant Graphics for Data Analysis. (Springer-Verlag New York, 2016).

75. Pedersen, T. L. ggraph: An Implementation of Grammar of Graphics for Graphs and Networks. Preprint at (2021).

76. Wilke, C. O. cowplot: Streamlined Plot Theme and Plot Annotations for ‘ggplot2’. Preprint at (2020).

77. Dowle, M. & Srinivasan, A. data.table: Extension of ‘data.frame’. Preprint at (2021).

78. Corporation, M. & Weston, S. doSNOW: Foreach Parallel Adaptor for the ‘snow’ Package. Preprint at (2020).

79. Microsoft & Weston, S. foreach: Provides Foreach Looping Construct. Preprint at (2020).

80. Lawrence, M. et al. Software for Computing and Annotating Genomic Ranges. PLoS Comput. Biol. 9, e1003118 (2013).

81. Clarke, E. & Sherrill-Mix, S. ggbeeswarm: Categorical Scatter (Violin Point) Plots. Preprint at (2017).

82. Kassambara, A. ggpubr: ‘ggplot2’ Based Publication Ready Plots. Preprint at (2020).

83. Warnes, G. R., Bolker, B. & Lumley, T. gtools: Various R Programming Tools. Preprint at (2021).

84. Müller, K. here: A Simpler Way to Find Your Files. Preprint at (2020).

85. Firke, S. janitor: Simple Tools for Examining and Cleaning Dirty Data. Preprint at (2021).

86. Bulik-Sullivan, B. et al. An atlas of genetic correlations across human diseases and traits. Nat. Genet. 47, 1236–1241 (2015).

87. Schauberger, P. & Walker, A. openxlsx: Read, Write and Edit xlsx Files. Preprint at (2021).

88. Rinker, T. W. {qdapTools}: Tools to Accompany the qdap Package. Preprint at (2015).

89. Wickham, H. & Bryan, J. readxl: Read Excel Files. Preprint at (2019).

90. Lawrence, M., Gentleman, R. & Carey, V. rtracklayer: An R package for interfacing with genome browsers. Bioinformatics 25, 1841–1842 (2009).

91. Wickham, H. et al. Welcome to the Tidyverse. J. Open Source Softw. 4, 1686 (2019).

